# Rapid feedback on hospital onset SARS-CoV-2 infections combining epidemiological and sequencing data

**DOI:** 10.1101/2020.11.12.20230326

**Authors:** Oliver T Stirrup, Joseph Hughes, Matthew Parker, David G Partridge, James G Shepherd, James Blackstone, Francesc Coll, Alexander J Keeley, Benjamin B Lindsey, Aleksandra Marek, Christine Peters, Joshua B Singer, The COVID-19 Genomics UK (COG-UK) consortium, Asif Tamuri, Thushan I de Silva, Emma C Thomson, Judith Breuer

## Abstract

**Background:** Rapid identification and investigation of healthcare-associated infections (HCAIs) is important for suppression of SARS-CoV-2, but the infection source for hospital onset COVID-19 infections (HOCIs) cannot always be readily identified based only on epidemiological data. Viral sequencing data provides additional information regarding potential transmission clusters, but the low mutation rate of SARS-CoV-2 can make interpretation using standard phylogenetic methods difficult.

**Methods:** We developed a novel statistical method and sequence reporting tool (SRT) that combines epidemiological and sequence data in order to provide a rapid assessment of the probability of HCAI among HOCI cases (defined as first positive test >48 hours following admission) and to identify infections that could plausibly constitute outbreak events. The method is designed for prospective use, but was validated using retrospective datasets from hospitals in Glasgow and Sheffield collected February-May 2020.

**Results:** We analysed data from 326 HOCIs. Among HOCIs with time-from-admission ≥8 days the SRT algorithm identified close sequence matches from the same ward for 160/244 (65.6%) and in the remainder 68/84 (81.0%) had at least one similar sequence elsewhere in the hospital, resulting in high estimated probabilities of within-ward and within-hospital transmission. For HOCIs with time-from-admission 3-7 days, the SRT probability of healthcare acquisition was >0.5 in 33/82 (40.2%).

**Conclusions:** The methodology developed can provide rapid feedback on HOCIs that could be useful for infection prevention and control teams, and warrants further prospective evaluation. The integration of epidemiological and sequence data is important given the low mutation rate of SARS-CoV-2 and its variable incubation period.

## Introduction

Nosocomial transmission of SARS-CoV-2 presents a significant health risk to both vulnerable patients and to healthcare workers (HCWs)^[1-5]^. There is a variable incubation period, extending up to day 14 from exposure to the virus in symptomatic cases^[6]^. It is also known that transmission is possible from asymptomatic or presymptomatic carriers^[7-10]^, complicating identification of hospital-acquisition among hospital onset COVID-19 infections (HOCIs) and tracing of likely sources of infection.

There is now substantial evidence from retrospective studies that genome sequencing of epidemic viruses, together with standard infection prevention and control (IPC) practice, better excludes nosocomial transmissions and better identifies routes of transmission than IPC investigation alone^[11-13]^. The development of rapid sequencing methods capable of generating pathogen genomes within 24-48 hours has recently created the potential for clinical IPC decisions to be informed by genetic data in near-real-time^[14]^. Although SARS-CoV-2 has a low mutation rate, estimated at around 2 changes per genome per month^[15]^, sufficient viral diversity does now exist for viral sequences to provide information regarding potential transmission clusters^[16]^. However, phylogenetic methods alone cannot reliably identify linked infections or the direction of transmission, and the need for clinical teams to gather additional patient data presents challenges to the timely interpretation of SARS-CoV-2 sequence data.

To overcome these barriers, we have developed a sequence reporting tool (SRT) that integrates genomic and epidemiological data from HOCIs to rapidly identify closely matched sequences within the hospital and assign a probability estimate for nosocomial infection. The output report is designed for prospective use to reduce the delay from SARS-CoV-2 sequencing to application of insights generated to IPC practice. As such, the probability model on which it is based has minimal computational requirements or need for local tuning and checking of model parameters. The work was conducted as part of the COVID-19 Genomics (COG) UK initiative, which sequences large numbers of SARS-CoV-2 viruses from hospitals and the community across the UK^[17]^. Here we describe the performance of the SRT using COG-UK sequence data for HOCI cases collected from Glasgow and Sheffield between February and May 2020 and explore how it may have provided additional useful information for IPC investigations.

## Methods

The SRT methodology is applied to HOCI cases, defined here as inpatients with first positive SARS-CoV-2 test or symptom onset >48 hours after admission, who were not suspected of having COVID-19 at admission. The SRT algorithm returns probability estimates for healthcare-associated infection (HCAI) in each HOCI case, i.e. the probability that they acquired their infection post-admission within the hospital, with information provided on closely matching viral sequences from the ward location at sampling and wider hospital. Results for individual HOCIs are evaluated in relation to the IPC classification system recommended by PHE and other UK public health bodies, based on interval from admission to positive test: 3-7 days post admission = indeterminate HCAI; 8-14 days post admission = probable HCAI; >14 days post admission = definite HCAI^[18]^. We also applied the PHE definition of healthcare-associated COVID-19 outbreaks^[18]^ (i.e. ≥2 cases associated with a specific ward, with at least one being a probable or definite HCAI) to ward-level data, and within each PHE-defined outbreak event we evaluated whether there was one or more clearly distinct genetic cluster. This was determined by consecutive linkage of each HOCI into clusters using a 2 SNP threshold (with HOCIs assigned to a genetic cluster if they were a sequence match to any member). Sequences with <90% genomic coverage were excluded from all analyses.

### Data collection and processing

Research Ethics for COG-UK was granted by the PHE Research Ethics and Governance group as part of the emergency response to COVID-19 (24 April 2020, REF: R&D NR0195).

#### Glasgow

During the first wave of SARS-CoV-2, the MRC-University of Glasgow Centre for Virus Research collected residual clinical samples from SARS-CoV-2 infected individuals following diagnosis at the West of Scotland Specialist Virology Centre. Samples were triaged for rapid sequencing using Oxford Nanopore Technologies (ONT) for suspected healthcare related infections or Illumina sequencing in all other cases (details in supplementary Appendix).

#### Sheffield

Residual clinical samples from SARS-CoV-2 positive cases diagnosed at Sheffield Teaching Hospitals NHS Foundation Trust were sequenced at the University of Sheffield using ARTIC network protocol^[19]^ and ONT. Throughout the epidemic, members of the IPC team were notified by the laboratory and by clinical teams of positive results and reviewed relevant areas to ensure optimisation of practice and appropriate management of patients. Electronic reports were created contemporaneously, including an assessment as to whether suspected linked cases were present based on ward level epidemiology. As part of SRT validation, these reports were accessed retrospectively by a study team member blind to the sequencing data and each included HOCI case was defined as being thought unlinked to other cases, a presumed index case in an outbreak or a presumed secondary case.

### HOCI classification algorithm

The sequence matching and probability score algorithm is run separately for each ‘focus sequence’ corresponding to a HOCI. We use associated metadata to assign other previously collected sequences to categories representing where the individual may be part of a SARS-COV-2 transmission network:

- Unit reference set: individual could be involved with transmission on same unit (ward/ICU etc) as focus sequence (look-back interval: 3 weeks)
- Institution reference set: individual could be involved with transmission in same institution/hospital as focus sequence (look-back interval: 3 weeks)
- Community reference set: individual could be involved with transmission outside of focus sequence institution (look-back interval: 6 weeks).

It is possible for samples to be members of multiple reference sets. For example an outpatient may be involved in SARS-CoV-2 transmission at the institution they attended and/or in community transmission.

For each run of the algorithm, pairwise comparisons are conducted between the focus sequence and each sequence within the unit reference set, institution reference set and community reference set. A reference set sequence is considered a close match to the focus sequence if there is a maximum of two SNP differences between them. This choice was based on reported healthcare-associated outbreak events^[14, 20]^ and the overall mutation rate of SARS-CoV-2 (details in supplementary Appendix).

#### Probability calculations

We use an expression of Bayes theorem to estimate probabilities for post-admission infection of each focus case divided by exposure on the unit, within the rest of the institution and from visitors (if they were allowed). An estimate of the prior probability (*P*_*prior*_) of post-admission infection for each focus case is modified to a posterior probability according to information provided by the sequence data. The algorithm is based on sound statistical principles, but involves heuristic approximations.

In symptomatic focus cases we base *P*_*prior*_ on the time interval (*t*) from admission to date of symptom onset or first positive test (if date of symptom onset not recorded). We calculate *P*_*prior*_= *F*(*t*), where *F*() is the cumulative distribution function of incubation times^[6]^. The derivation of this method is given in the supplementary appendix.

In theory, it would be optimal to use all of the information in the *exact* sequences observed. However, with the goal of constructing a computationally simple algorithm, we base our calculations on the probability of observing a *similar* sequence (within 2 SNPs) to that actually observed for each focus case conditional on each potential infection source/location. We require *estimates* of the probability of observing a similar sequence to the focus sequence conditional on infection in the community, current unit/ward or elsewhere in the hospital/institution, or from a visitor. For the unit and hospital, we estimate this using the observed sequence match proportion (on pairwise comparison to the focus sequence) in the unit reference set and institution reference set, respectively. For community- or visitor-acquired infection we use a weighted proportion of matching sequences in the community reference set, with weightings determined by a calibration model that describes geographic clustering of similar sequences among community-acquired infections (described in supplementary appendix). The geographic weighting model was fitted separately for each study site using sequences strongly thought to represent community-acquired infection: all community-sampled sequences and patients presenting to the Emergency Department with COVID-19, excluding those recorded as being healthcare workers.

### Software

The analysis was conducted in R (v. 4.0.2, R Foundation, Vienna), using sequence processing and comparison functions from *ape* (v5.4) and geospatial functions in the *PostcodesioR* (v0.1.1) and *gmt* packages (v2,0). The algorithm has also been implemented as a standalone SRT for prospective use^[21]^ within COV-GLUE^[22]^.

## Results

### Study populations

#### Glasgow

The Glasgow dataset included 1199 viral sequences (available as of 23^rd^ June 2020): 426 of which were derived from community sampling sites, 351 from patients presenting to the Emergency Department or hospital assessment locations such as acute medical units, 398 from hospital inpatients and 24 from outpatients. Limited data were available regarding the total number of HCWs testing positive and identification of their samples amongst those from the community-sampling sites, but 15 sequences were recorded as having been obtained from HCWs. The first positive test dates ranged from 3rd March to 27th May 2020. All consensus sequences had genomic coverage >90%.

We applied the SRT algorithm to data from three hospitals with the required metadata available, for which 128/246 inpatient cases with sequences were HOCIs. Two of these patients had been transferred from another hospital within 14 days prior to their positive test and were not processed as focus sequences. One patient for whom we were unable to determine the exact sampling location within the hospital was also excluded, leaving 125 HOCI cases for analysis. Population sequencing coverage was 536/1578 (34.0%) overall for patients at the three hospitals and 128/328 (39.0%) for HOCIs specifically (Figure S1).

#### Sheffield

The Sheffield dataset included 1630 viral sequences with accompanying metadata (available as of 10^th^ October 2020): 714 were from inpatients, 117 were from outpatients and 799 were from HCWs. For the purpose of the retrospective evaluation, the 447/714 inpatient samples taken on date of admission were assumed to represent community-onset cases and used to calibrate the model. The first positive test dates ranged from 23rd February to 30th May 2020. One sequence with genome coverage <90% was dropped from further analysis (from an inpatient on date of admission). 201 of the inpatients were HOCIs. Population sequencing coverage was 714/977 (73.1%) overall for inpatients, 201/261 (77.0%) for HOCIs specifically and 799/962 (83.1%) for HCWs.

### Comparison to standard PHE classification

The SRT algorithm results in comparison to standard PHE classifications are summarised in Figure 1 and Table 1. The majority of HOCI cases in Glasgow (78/125, 62.4%) and over a third of those in Sheffield (71/201, 35.3%) met the PHE definition of a definite HCAI and so are known to have acquired the virus post-admission irrespective of sequencing results. The probable HCAI cases formed the next largest group at each site. Overall, the SRT algorithm identified close sequence matches from the same ward for 66.4% of definite and 64.2% of probable HCAIs, indicating likely within-ward transmission (examples provided in Case Studies 1-3). When one or more close sequence matches was identified on the ward of the focus sequence, the SRT probability of infection on the ward was >0.5 in 185/189 cases (Figure 2). For indeterminate HCAIs the SRT probability of HCAI was >0.5 in 33/82 (40.2%), and in 27/33 (81.8%) cases a close sequence match on the ward was present. Overall, 14/125 (11.2%) HOCIs in Glasgow and 175/201 (87.1%) in Sheffield had at least one close sequence match to a HCW sample, reflecting the much greater availability of sequences from HCWs in the Sheffield dataset.

**Table 1.**
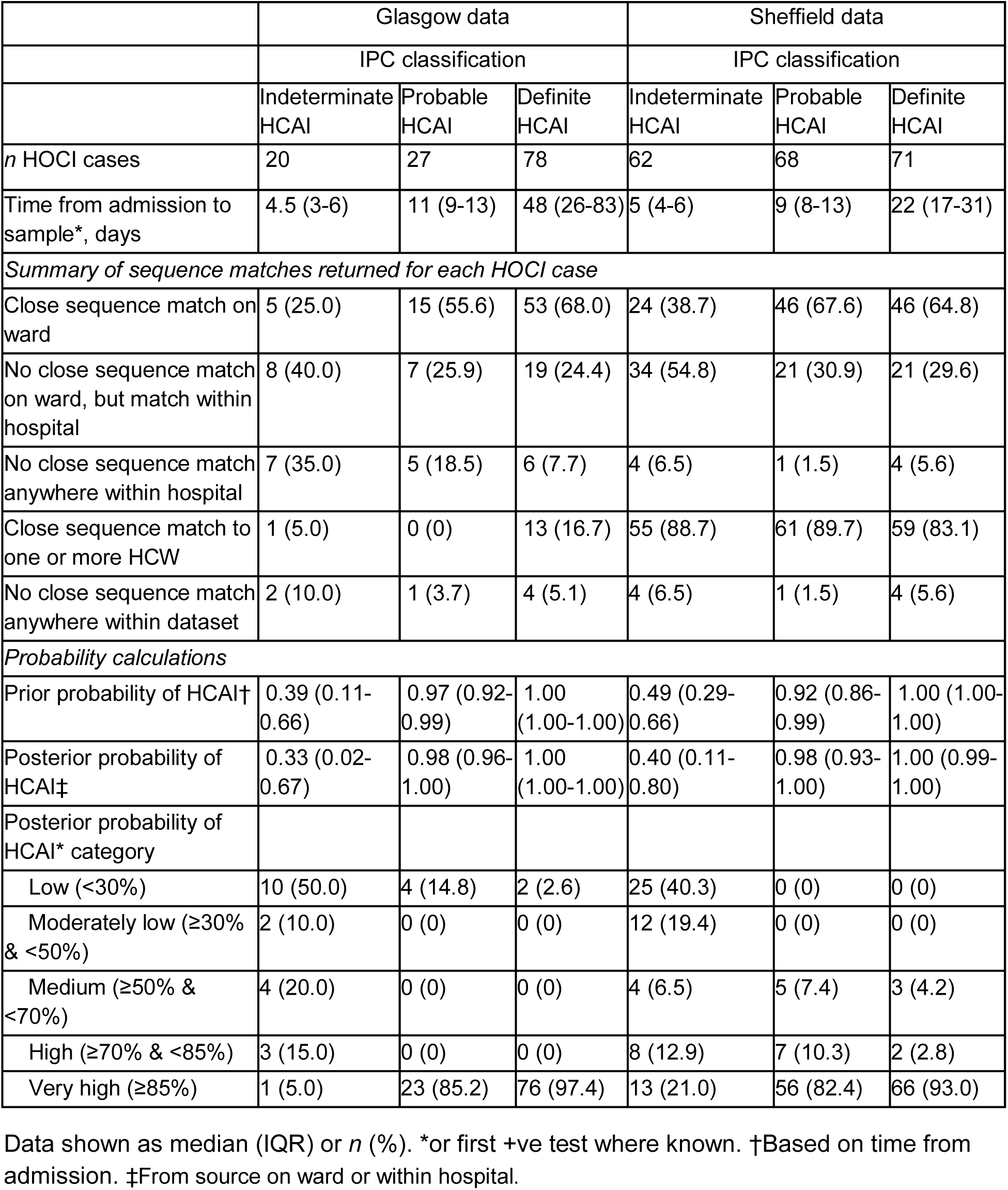
Summary of sequence reporting tool outputs for the Glasgow and Sheffield datasets, according to standard IPC definitions recommended by PHE regarding likelihood of healthcare-associated infection (HCAI)

|  | Glasgow data |  |  | Sheffield data |  |  |
| --- | --- | --- | --- | --- | --- | --- |
|  | IPC classification |  |  | IPC classification |  |  |
|  | Indeterminate HCAI | Probable HCAI | Definite HCAI | Indeterminate HCAI | Probable HCAI | Definite HCAI |
| *n* HOCI cases | 20 | 27 | 78 | 62 | 68 | 71 |
| Time from admission to sample*, days | 4.5 (3-6) | 11 (9-13) | 48 (26-83) | 5 (4-6) | 9 (8-13) | 22 (17-31) |
| *Summary of sequence matches returned for each HOCI case* |  |  |  |  |  |  |
| Close sequence match on ward | 5 (25.0) | 15 (55.6) | 53 (68.0) | 24 (38.7) | 46 (67.6) | 46 (64.8) |
| No close sequence match on ward, but match within hospital | 8 (40.0) | 7 (25.9) | 19 (24.4) | 34 (54.8) | 21 (30.9) | 21 (29.6) |
| No close sequence match anywhere within hospital | 7 (35.0) | 5 (18.5) | 6 (7.7) | 4 (6.5) | 1 (1.5) | 4 (5.6) |
| Close sequence match to one or more HCW | 1 (5.0) | 0 (0) | 13 (16.7) | 55 (88.7) | 61 (89.7) | 59 (83.1) |
| No close sequence match anywhere within dataset | 2 (10.0) | 1 (3.7) | 4 (5.1) | 4 (6.5) | 1 (1.5) | 4 (5.6) |
| *Probability calculations* |  |  |  |  |  |  |
| Prior probability of HCAI† | 0.39 (0.11-0.66) | 0.97 (0.92-0.99) | 1.00 (1.00-1.00) | 0.49 (0.29-0.66) | 0.92 (0.86-0.99) | 1.00 (1.00-1.00) |
| Posterior probability of HCAI‡ | 0.33 (0.02-0.67) | 0.98 (0.96-1.00) | 1.00 (1.00-1.00) | 0.40 (0.11-0.80) | 0.98 (0.93-1.00) | 1.00 (0.99-1.00) |
| Posterior probability of HCAI* category |  |  |  |  |  |  |
| Low (<30%) | 10 (50.0) | 4 (14.8) | 2 (2.6) | 25 (40.3) | 0 (0) | 0 (0) |
| Moderately low (≥30% & <50%) | 2 (10.0) | 0 (0) | 0 (0) | 12 (19.4) | 0 (0) | 0 (0) |
| Medium (≥50% & <70%) | 4 (20.0) | 0 (0) | 0 (0) | 4 (6.5) | 5 (7.4) | 3 (4.2) |
| High (≥70% & <85%) | 3 (15.0) | 0 (0) | 0 (0) | 8 (12.9) | 7 (10.3) | 2 (2.8) |
| Very high (≥85%) | 1 (5.0) | 23 (85.2) | 76 (97.4) | 13 (21.0) | 56 (82.4) | 66 (93.0) |

Data shown as median (IQR) or *n* (%). \*or first +ve test where known. †Based on time from admission. ‡From source on ward or within hospital.

**Figure 1.**
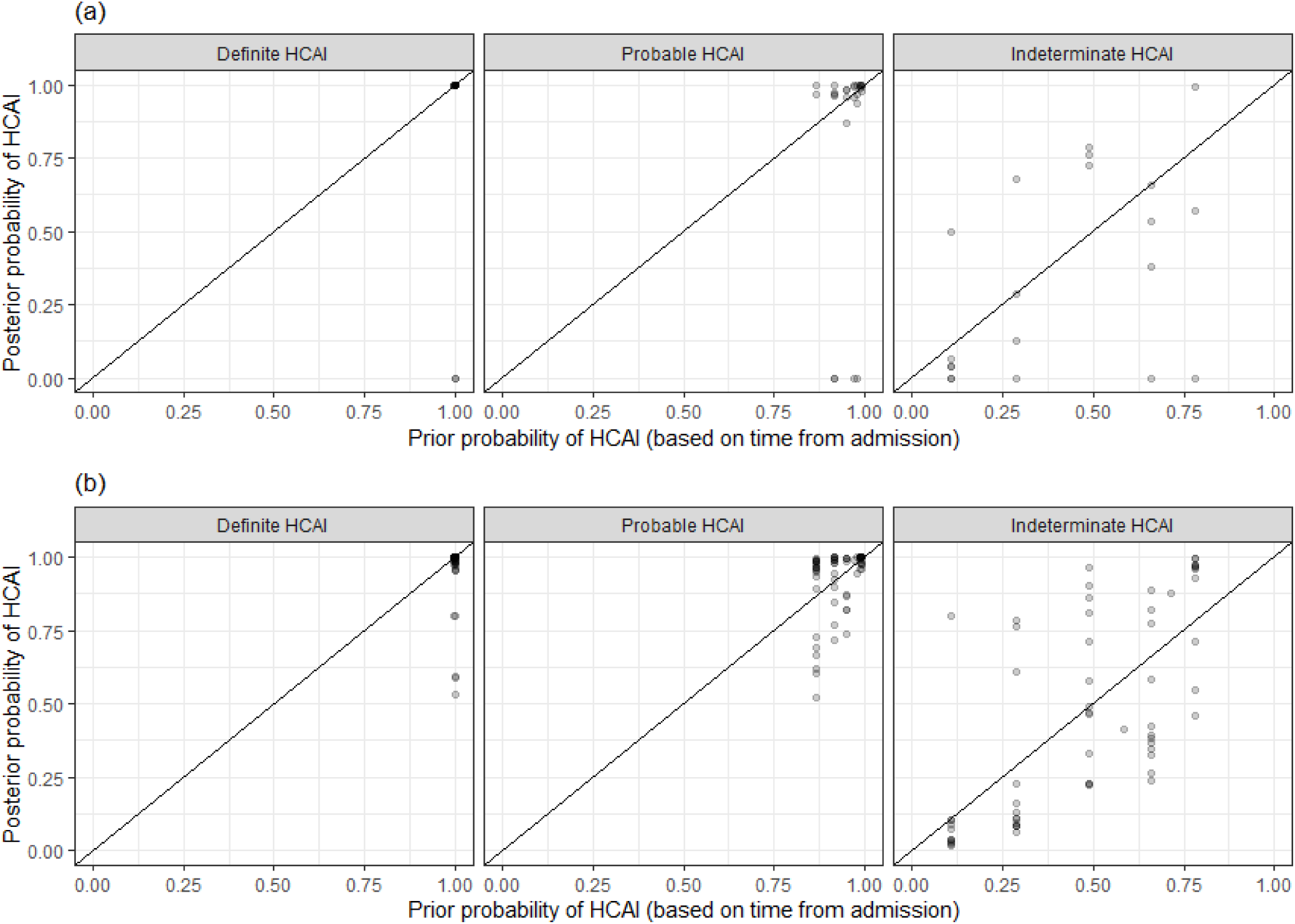
Plot of the posterior probability of healthcare-associated infection (HCAI) for (a) Glasgow and (b) Sheffield HOCIs from the sequence reporting tool algorithm against the prior probability of HCAI based only on time from admission to diagnosis, grouped by standard IPC classification recommended by PHE.

**Figure 2.**
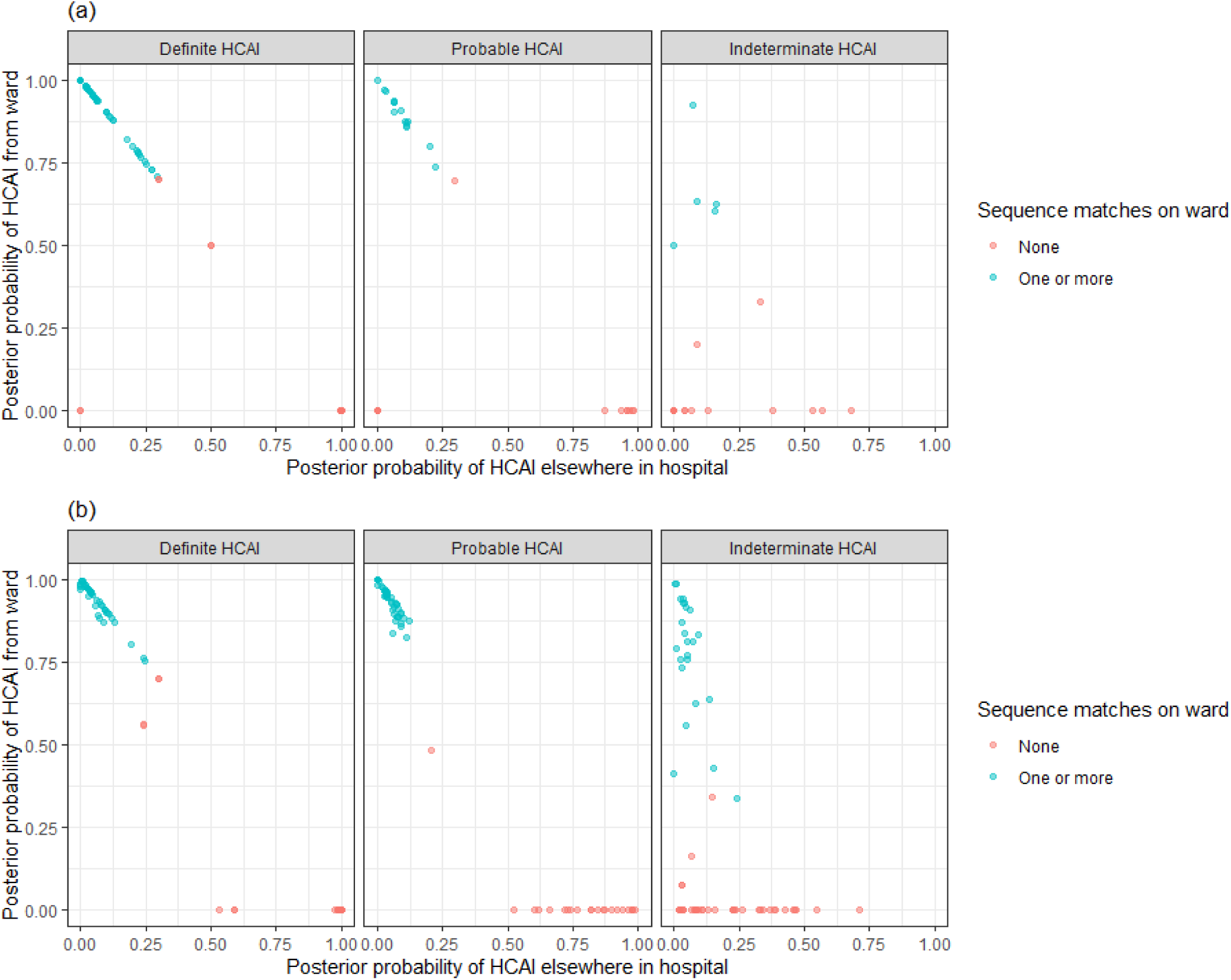
Plot of the posterior probabilities of healthcare-associated infection (HCAI) estimated using the sequence reporting tool algorithm from a source on the current ward versus a source elsewhere in the hospital for (a) Glasgow and (b) Sheffield HOCIs grouped by standard PHE classification. In cases where there are no close sequence matches in the dataset (including among community cases), the results returned are based solely on the priors and the metadata; this explains the fact that there are some cases with estimated posterior probability of infection on the ward greater than 0.5 for whom there were no sequence matches on the ward.

In 16/244 (6.6%) cases that met the probable or definite HCAI definitions, there was no sequence match within the hospital; this is likely due to incomplete sequence data from SARS-CoV-2 hospitalised cases and staff (with population sequencing coverage <40% patients and very limited for staff from Glasgow and ≈75% of patients and staff in Sheffield) and the presence of undiagnosed carriers. To reflect this we designed the SRT report to return the following message in such situations “This is a probable/definite HCAI based on admission date, but we have not found genetic evidence of transmission within the hospital”. There were 26 HOCIs in the Sheffield dataset for whom it was recorded that visitors were allowed on the ward at time of sampling. In three of these the estimated probability of infection from a visitor was between 0.4 and 0.5 (all had ≥18 days from admission to diagnosis and no close sequence matches on the ward).

Within the Sheffield dataset we identified six wards with two genetically distinct outbreak clusters (of two or more patients) and three wards with three distinct outbreaks (see Case Study 3). Standard IPC assessment had classified each as a single outbreak. We also identified 10 and 44 HOCIs in the Glasgow and Sheffield datasets, respectively, with no apparent genetic linkage to other HOCI cases on the ward but who met the PHE definition of inclusion within an outbreak event (Table 2). There were two HOCIs in the Sheffield dataset which showed a close sequence match to another case on the same ward with interval from admission to sample date ≤2 days.

**Table 2.** Summary of distinct outbreak events for the Glasgow and Sheffield datasets, according to standard PHE definition and with the addition of sequence data

|  | Glasgow data | Sheffield data |
| --- | --- | --- |
| *n* HOCl cases | 125 | 201 |
| *n* ward locations | 44 | 38 |
| *Sequence matches per HOCl case* |  |  |
| *n* sequence matches from same ward, median (IQR, range) | 1 (0-5, 0-12) | 1 (0-4, 0-18) |
| *n* sequence matches from rest of hospital, median (IQR, range) | 3 (1-8, 0-52) | 27 (5-52, 0-150) |
| *Standard PHE definition of outbreak event* |  |  |
| HOCl cases part of ward outbreak event, *n* (%) | 95 (76.0) | 184 (91.5) |
| *n* ward outbreak events | 17 | 24 |
| *n* HOCl cases per ward outbreak event, median (IQR, range) | 4 (2-8, 2-17) | 5 (3.5-10.5, 2-28) |
| Days from first to last case in outbreak, median (IQR, range) | 8 (6-15, 0-31 ) | 18 (13-34, 3-68) |
| *n* wards with more than one distinct outbreak event | 0 | 0 |
| *Outbreak events with sequence linkage* |  |  |
| HOCl cases part of ward outbreak event, *n* (%) | 85 (68.0) | 140 (69.7) |
| *n* ward outbreak events | 16 | 33 |
| *n* HOCl cases per ward outbreak event, median (IQR, range) | 3.5 (2-8, 2-16) | 3 (2-4, 1-19) |
| Days from first to last case in outbreak, median (IQR, range) | 6 (4-9, 0-15) | 4 (2-8, 0-17) |
| *n* wards with more than one distinct outbreak event | 0 | 9* |

\*In three wards there were three genetically distinct outbreak events.

### Comparison to local IPC conclusions in Sheffield

Detailed contemporaneous notes collected by IPC teams in Sheffield classified 18/201 HOCIs as the index case in outbreaks. IPC staff defined an index case as the first detected in an environment regardless of prior inpatient stay and, correspondingly, of these 14/18 were the first sequence on their ward and one was the second (the first 1 day earlier from a different bay on the ward was also recorded as an index case, and IPC staff deemed a ward outbreak with unclear index or possibly 2 index cases). Of the 18 index cases 11 showed at least one subsequent close sequence match on the same ward (the 2 index cases on a single ward were not genetically similar, and for 1/18 there were no subsequent sequences from the ward). The median SRT probability of HCAI was 0.70 (IQR 0.22-1.00, range 0.04-1, >0.5 in 12/18).

A further 144/201 HOCIs were classified as being part of local outbreaks, and among these the median SRT probability of HCAI was 0.98 (IQR 0.89-1.00; range 0.02-1.00; >0.5 in 129/144) with one or more close sequence match on the same ward in 104/144. The remaining 39/201 HOCIs, including 10 that were not recorded as HOCIs at the time, were classified by the IPC teams as not being part of local outbreaks). Among these the median SRT probability of HCAI was 0.74 (IQR 0.23-0.99, range 0.02-1.00; >0.5 in 23/39), with one or more close sequence matches on the same ward in 7/39.

#### Case Study 1

Figure 3 shows a phylogenetic tree of eight HOCIs within a single ward at a Glasgow hospital (Hospital 5, Unit 93), alongside associated meta-data and SRT probability outputs. The first HOCI detected (UID0032) was transferred from another hospital within the previous 2 weeks and so SRT output was not generated. All subsequent HOCIs return close sequence matches to at least one prior case on the ward, leading to SRT probability estimates of ward-acquired infection >0.9, even for UID0017 (an indeterminate HCAI). The phylogenetic tree indicates UID0032 has a SNP lacked by most of the cases identified on the ward, and therefore did not seed all of the cases in the outbreak cluster. Also shown on the tree is a single HOCI from a different ward in the same hospital (UID0025); this individual was an indeterminate HCAI, but a higher proportion of similar viral sequences within the hospital in comparison to their local community led to a SRT result of probable hospital-acquired infection.

**Figure 3.**
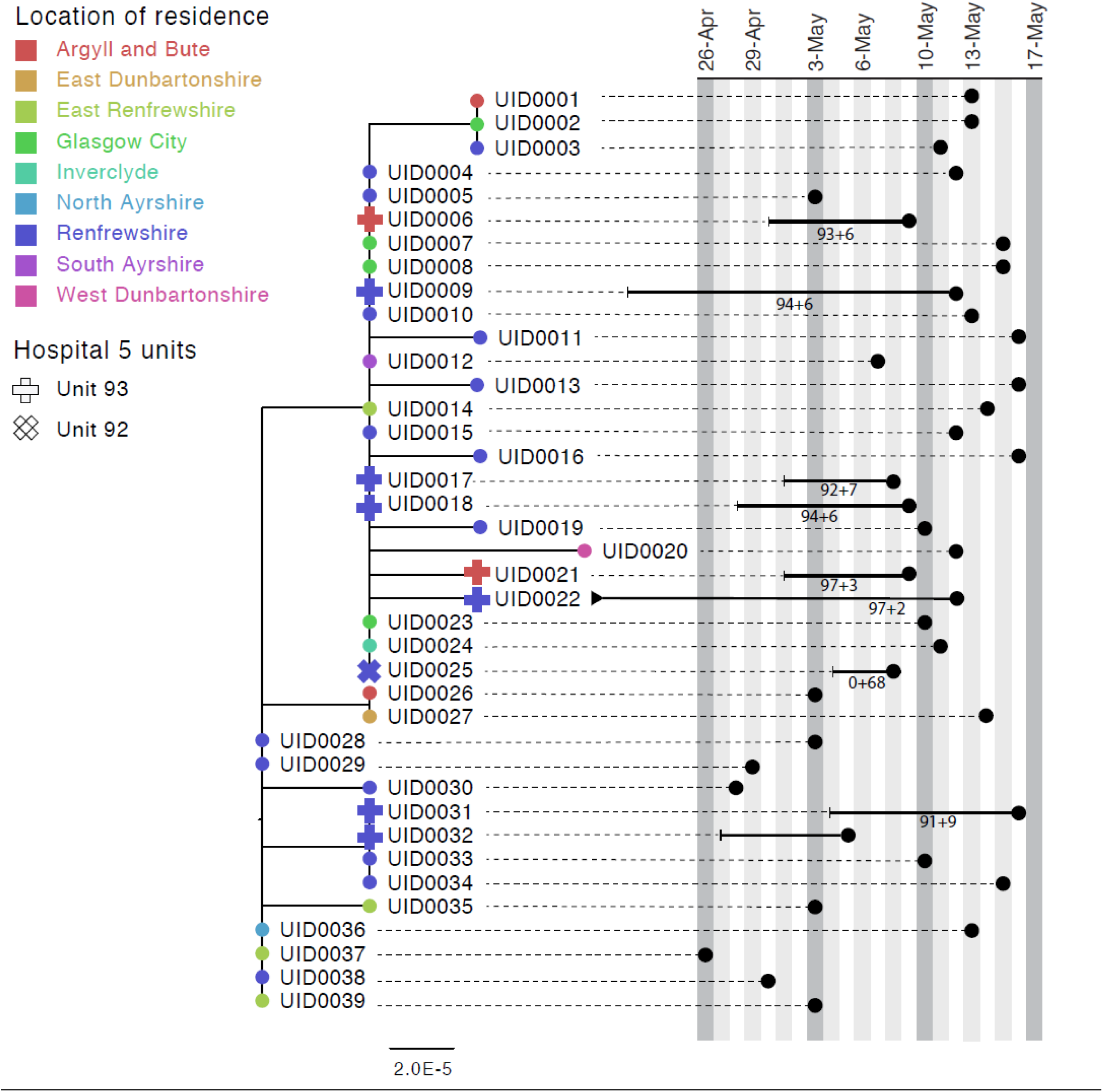
Maximum-likelihood phylogeny of the sequences found in Hospital 5 Unit 93 and Unit 92 up until the 16th of May of the Glasgow dataset. The black lines represent the time from admission to sampling. The values below the line are the posterior probability for unit infection + the posterior probability of hospital infection from the sequence reporting tool. The tip nodes are coloured according to the local authority area of the community surveillance sequences (circles) or of the patients (crosses).

#### Case Study 2

Figure 4 shows a phylogenetic tree indicating complex transmission networks across multiple hospitals in the Glasgow area (with SRT outputs for Hospitals 2 and 4). A monophyletic cluster of HOCIs can be seen in Hospital 2 Unit 48, with the first detected case identified by the SRT as a hospital-acquired and the rest unit-acquired infections. A paraphyletic group of HOCIs was detected in Hospital 4 Unit 69. Patient 1 (UID0042) was screened for COVID in Unit 69 on 14.04.20 after developing a cough and oxygen requirement. The patient was moved from the nightingale area to a single room on the ward on 14.04.20 and was confirmed positive on 15.04.20.

**Figure 4.**
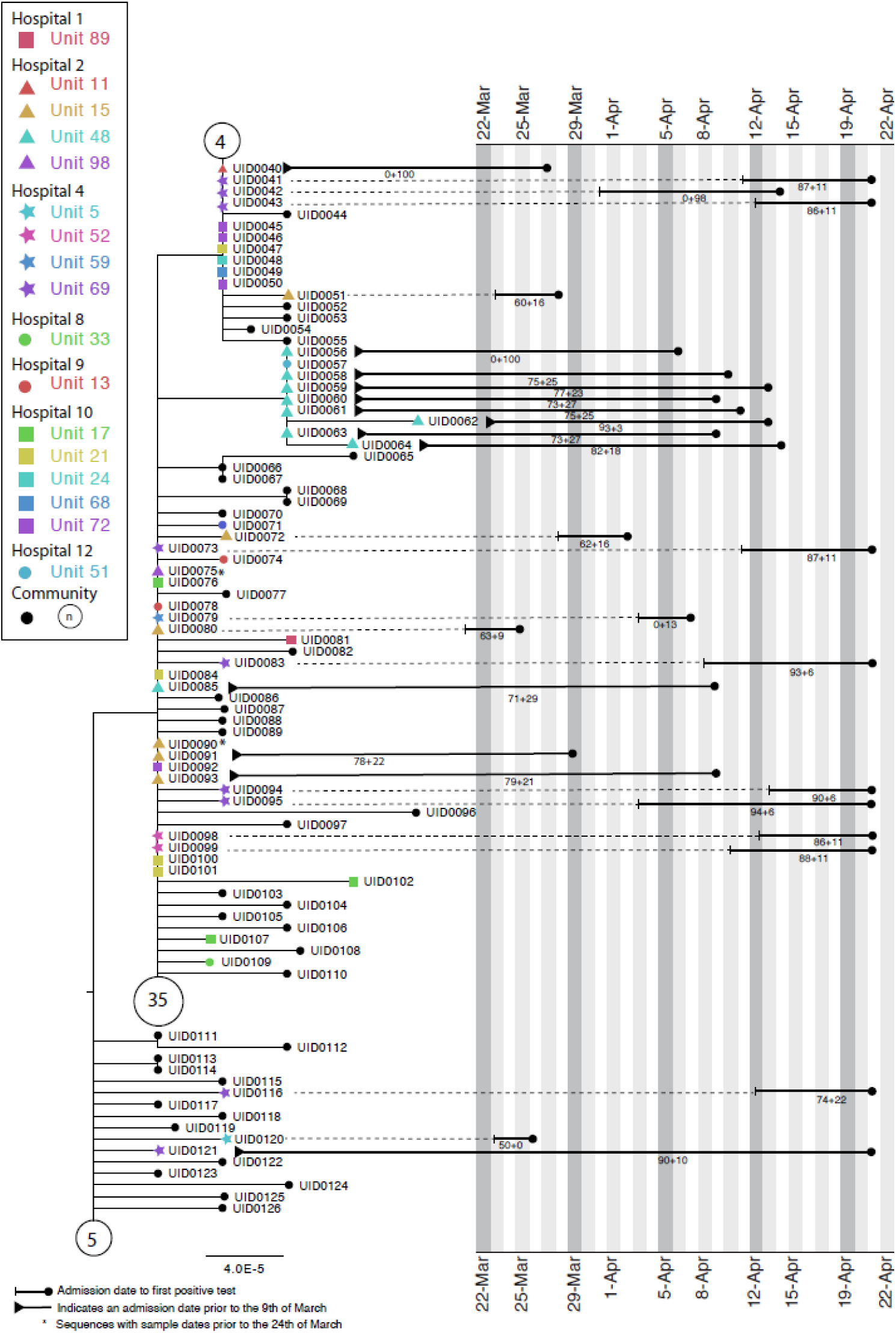
Maximum-likelihood tree for sequences found in Hospital 2 Unit 48 and Hospital 4 Unit 69 of the Glasgow dataset up until the 21st of April (inclusive). The circles with numbers represent the number of community sequences that are identical and at the base of each lineage (n=5, n=35, n=4). Tree tips with black circles represent further community sequences. The black lines represent the time from admission to sampling. The values below the line are the posterior probability for unit infection + the posterior probability of hospital infection from the sequence reporting tool.

On 20.04.20 a second patient on Unit 69 (not sequenced) was screened after developing a cough and pyrexia and confirmed positive on 21.04.2020. The patient was in a single room at the time of symptom onset, however they had been in the main nightingale ward opposite patient 1 for 5 days. At this point 13 asymptomatic contacts in Unit 69 were screened, and 8 (UID0043, UID0073, UID0041, UID0095, UID0116, UID0094, UID0083, UID0121) were positive.

These cases are all identified as hospital-acquired or unit-acquired infections and can be grouped into a genetically similar cluster with a maximum pairwise distance of 2 SNPs between each member and its nearest neighbour. However, this cluster clearly represents multiple introductions of SARS-CoV-2 onto the ward.

#### Case Study 3

Figure 5 shows phylogenetic trees relating to three distinct viral lineages identified on a single ward in the Sheffield dataset (classified by contemporaneous IPC investigation as a single outbreak). Two of these lineages also include sequences from inpatients sampled from other wards within the same hospital. Detailed ward movement data highlighted additional possible links between patients in the B.2.1 cluster. Both UID0149 and UID0157 were present at LOC0111 prior to their sample dates.

**Figure 5.**
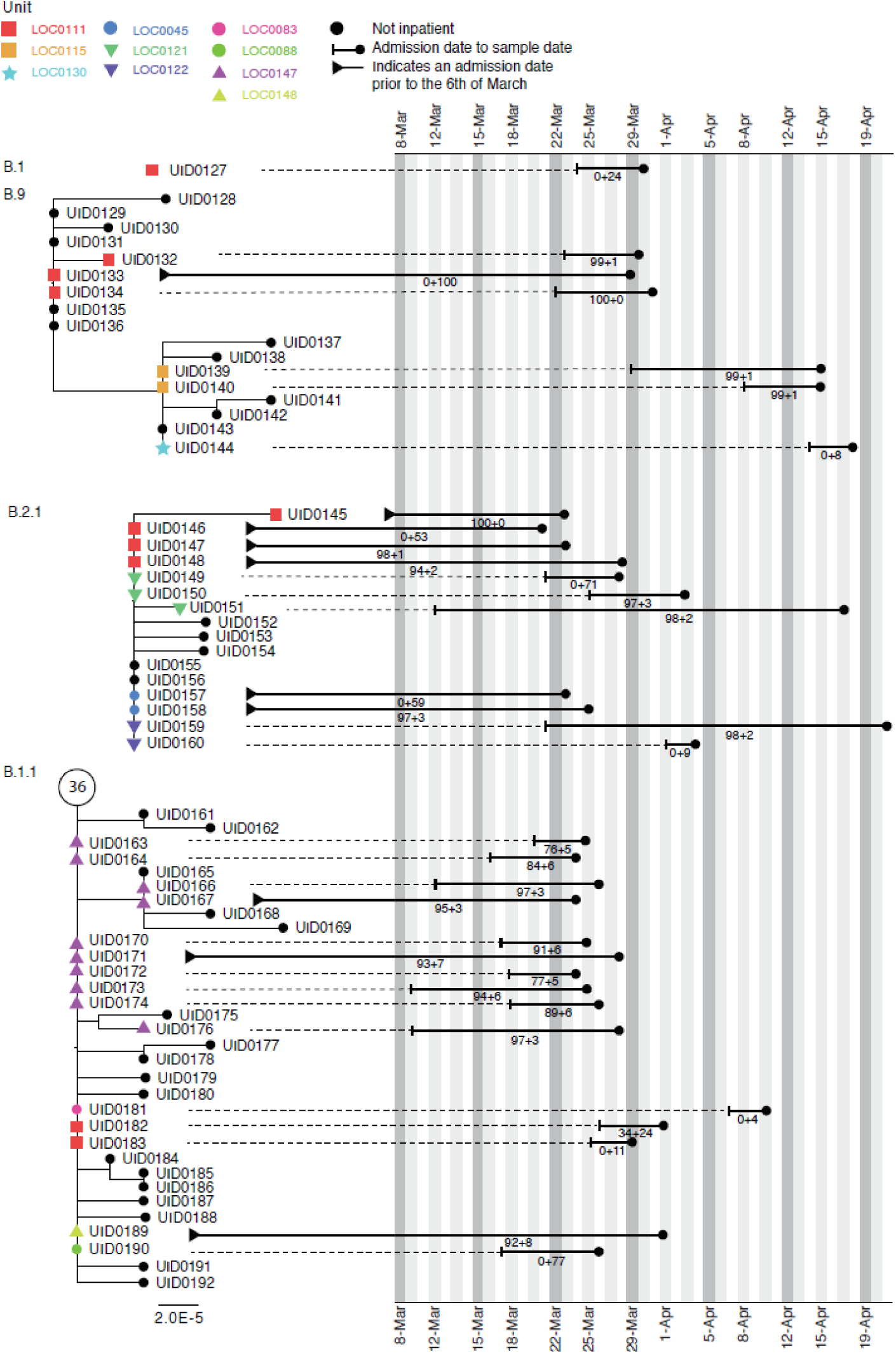
Maximum-likelihood phylogeny of the sequences found in Location ‘0111’ in the Sheffield dataset, also including patients at several other ward locations. The tree tip nodes are coloured according to ward locations. The black lines represent the time from admission to sampling. The values below the line are the posterior probability for unit infection + the posterior probability of hospital infection from the sequence reporting tool.

## Discussion

We have developed a novel approach for identification and investigation of hospital-acquired SARS-CoV-2 infections combining epidemiological and sequencing data, designed to provide rapid and concise feedback to IPC teams working to prevent nosocomial transmission. Through retrospective application to clinical datasets, we have demonstrated that the methodology is able to provide confirmatory evidence for most PHE-defined definite and probable HCAIs and provide further information regarding indeterminate HCAIs. Thus the SRT may allow IPC teams to optimise their use of resources, concentrating audit, review and educational interventions on areas with likely nosocomial acquisition events.

While the SRT is not likely to change IPC conclusions in cases meeting the definition of ‘definite’ or ‘probable’ HCAI based on interval from admission to symptom onset, in 91% of cases it did identify patients in the same ward or elsewhere in the hospital who could plausibly be linked to the HOCI within a single outbreak event. Those definite and probable HOCIs without close sequence matches are likely to reflect transmission from sources within the hospital that have either not been diagnosed or who were diagnosed without viral sequencing. In such cases it is impossible to calculate a probability of transmission and the SRT will simply state that no sequence matches were found within the hospital.

For cases meeting the definition of ‘indeterminate healthcare associated’, the probability scores returned would be useful for IPC teams. These probabilities are dependent on comparison to sequences from cases of community-acquired infection obtained either from direct community sampling or from patients sampled at admission. The Sheffield dataset was lacking the former data source, but the SRT nonetheless classified a similar proportion of ‘indeterminate healthcare associated’ HOCIs as community-acquired infections to that found in the Glasgow dataset (approximately 60%).

Current PHE guidelines define healthcare-associated COVID-19 outbreaks as two or more cases associated with a specific setting (e.g. ward), with at least one case having illness onset after 8 days of admission^[18]^. However, the guidelines note that “investigations of healthcare associated SARS-CoV-2 infection should also take into account COVID-19 cases categorised as ‘indeterminate healthcare associated’ (i.e. onset 3-7 days after admission), for which our SRT output would be useful. In most HOCIs meeting this definition of inclusion within an outbreak event, we found evidence of clusters of similar viral sequences located on the ward concerned, and the SRT results were in line with available local IPC classifications in the majority of cases. However, a substantial minority (54/279) of HOCIs although assumed to be part of a ward outbreak, were, in fact, isolated cases for which the sequencing data refuted genetic linkage to other sequences from the ward. The SRT also provided evidence of wards where IPC-defined outbreak events comprised two or three clearly distinct viral lineages (Case Report 3).

The retrospective datasets analysed in this study represent the first few months of the COVID-19 epidemic in the UK, and nosocomial transmission of the virus in the UK during this period has previously been reported at multiple sites^[14, 23, 24]^. HCWs were at increased risk of infection and adverse health outcomes^[1, 2, 4, 5, 25]^ and could have been important drivers of nosocomial transmission^[8]^. Data were limited for Glasgow but the Sheffield dataset contained a large number of sequences obtained from HCWs, with population sequencing coverage for this group >80%, and there was a close sequence match to at least one HCW observed for 87% of HOCIs. Our analysis has not evaluated direction of transmission to or from HCWs, but they were clearly linked into transmission networks within the hospital. A limitation of the current SRT approach and of the retrospective data available is that they do not include detailed information regarding work locations for HCWs. However, prospective use of the SRT would allow IPC teams to investigate linkage from a HOCI to any HCWs flagged as having a close sequence match.

While a phylogenetic approach is useful in excluding direct transmission between cases, it can be more problematic to confirm transmission source^[26]^. Phylogenetic models can evaluate the full genetic information provided by viral sequence data, but there are challenges in incorporating and summarising associated patient meta-data in a timely fashion^[27]^. There will be cases in which phylogenetic analysis would provide information beyond that returned by the SRT. However, fully integrated epidemiological and phylogenetic analysis of hospital outbreaks is resource-intensive, presenting challenges in delivering the rapid turnaround and scale-up required to provide clear feedback to hospital IPC teams outside of research-intensive settings.

Comparison of SRT output to phylogenetic trees in a number of test cases suggested that some clusters of genetically similar cases identified within a specific ward likely represented more than one transmission event onto the ward from similar viral lineages circulating within the healthcare system. Whilst monophyletic clusters associated with a single location are easier to interpret, we consider the presence of viruses within a ward or hospital that are genetically similar to a HOCI as evidence for nosocomial infection even when they are not plausible transmission sources themselves, given the potential for asymptomatic transmission^[7-10]^ and complex transmission networks^[14]^.

The SRT uses a number of heuristic approximations in order to provide an integrated summary of epidemiological and sequence data. However, this choice is associated with the limitation that it does not provide a full probabilistic model of potential transmission networks. Further development of the SRT would also aim to more fully incorporate patient movement data and shift locations for HCWs.

Our novel approach to the investigation of HOCIs has shown promising characteristics on retrospective application to two clinical datasets. The SRT described allows rapid feedback on HOCIs that integrates epidemiological and sequencing data to generate a simplified report at the time that sequence data become available. Its prospective use will be evaluated in a multicentre trial in late 2020 and early 2021. The methodology has been developed for hospital inpatients, but the principles may also be applicable to other settings.

## Data Availability

The sequence data analysed are included within publicly available datasets. However, due to data governance restrictions it is not possible to openly share the associated meta-data for the analysis described.

https://www.cogconsortium.uk/data/

## Acknowledgements

COG-UK HOCI is funded by the COG-UK consortium, which is supported by funding from the Medical Research Council (MRC) part of UK Research & Innovation (UKRI), the National Institute of Health Research (NIHR) and Genome Research Limited, operating as the Wellcome Sanger Institute. JBr receives funding from the NIHR ULC/UCLH Biomedical Research Centre. FC is funded by Wellcome (grant number: 201344/Z/16/Z). MDP is funded by the NIHR Sheffield Biomedical Research Centre (BRC - IS-BRC-1215-20017). We acknowledge the help of the UCL Comprehensive Clinical Trials Unit. The authors wish to thank the NHS Greater Glasgow and Clyde and Sheffield Teaching Hospitals NHS Foundation Trust infection prevention and control teams for provision of data. The authors thank Michael Chapman for his assistance in the development of this project.

## Online-only Supplementary Appendix

### Methods

#### Details of sequencing protocols

##### Glasgow

Sequencing with ONT followed the protocols developed by the ARTIC network (v1 and v2) https://artic.network/ncov-2019. The reads were aligned to the reference strain (MN908947) using minimap2 (https://doi.org/10.1093/bioinformatics/bty191) and denoised using nanopolish (https://www.nature.com/articles/nmeth.3444) prior to primer trimming and consensus calling with iVar using a minimum depth of 20 reads (https://doi.org/10.1186/s13059-018-1618-7). Sequencing with Illumina also used the ARTIC network protocol for amplicon generation but was followed by a DNA KAPA library preparation kit (Roche) and indexing with NEBNext multiplex oligos (NEB) using 7 PCR cycles. Libraries were pooled and loaded on a MiSeqV2 cartridge. Illumina reads were processed with the PrimalAlign pipeline (https://github.com/rjorton/PrimalAlign). Briefly, reads were trimmed using trim_galore (https://www.bioinformatics.babraham.ac.uk/projects/trim_galore/) aligned to the reference using BWA (10.1093/bioinformatics/btp698). Then, amplicon primers were removed and the consensus called with a read depth of 10 using iVar (https://doi.org/10.1186/s13059-018-1618-7). Metadata associated with each sample was collated in a redcap database (https://www.project-redcap.org/).

##### Sheffield

Sequencing with ONT followed the protocols developed by the ARTIC network (v1 and v2) https://artic.network/ncov-2019. Following base calling, data were demultiplexed using ONT Guppy using a high accuracy model. Reads were filtered based on quality and length (400 to 700bp), then mapped to the Wuhan reference genome and primer sites trimmed. Reads were then downsampled to 200x coverage in each direction. Variants were called using nanopolish (https://github.com/jts/nanopolish) and used to determine changes from the reference. Consensus sequences were constructed using reference and variants called.

#### Further details of reference set definitions

##### Data sources for algorithm

There are two potential sources of data for the HOCI classification algorithm. Firstly, there are *institution-sampled sequences*: these include all viral sequences from samples obtained within the institution/hospital. These sequences are linked to meta-data providing basic information regarding the patient concerned and details of the sample from which the sequence was obtained. Secondly, there are *community-sampled sequences*: these include all relevant sequences obtained from samples from testing within the local community. These sequences are associated with a more limited set of linked meta-data describing date of sample, residential outer postcode of subject and place of work if they are recorded as being a HCW.

##### Unit reference set

This data set comprises all institution-sampled sequences sampled on or ≤3 weeks prior to (or ≤2 days after for the prospective version of the SRT) the sample date of the focus sequence and for which both the institution and the unit is the same as that for the focus sequence.

##### Institution reference set

This data set comprises firstly all institution-sampled sequences from HCWs, outpatients and inpatients diagnosed >48 h after admission for which the institution matches that of the focus sequence sampled on or ≤3 weeks prior to (or ≤2 days after for the prospective version of the SRT) the sample date of the focus sequence and for which the unit is either not the same as that for the focus sequence or is missing. Secondly, the data set includes all institution-sampled sequences from A&E patients or inpatients diagnosed ≤2 days after admission for which the institutionID matches that of the focus sequence sampled between (inclusively) 3 weeks and 3 days prior to the sample date of the focus sequence and for which the unit is either not the same as that for the focus sequence or is missing. Thirdly, this data set also includes the subset of community-sampled sequences of healthcare workers at the same institution as the focus sequence.

##### Community reference set

This data set comprises firstly all community-sampled sequences sampled on or ≤6 weeks prior to (or ≤2 days after for the prospective version of the SRT) the sample date of the focus sequence. This data set also includes institution-sampled sequences sampled on or ≤6 weeks prior to (or ≤2 days after for the prospective version of the SRT) the sample date of the focus sequence from all non-inpatient samples, and those inpatients for whom sample date and symptom onset date (if recorded) are both ≤2 days after the admission date.

Note that some institution-sampled sequences will contribute to both the community reference set and either the unit reference set or the institution reference set (e.g. outpatients sampled within 3 weeks prior to the focus sequence would be included in both the community reference set and the institution reference set). HCWs recorded among the community-sampled sequences within ≤3 weeks prior to the sample date of the focus sequence will also be included in both the community reference set and the institution reference set if their workplace matches the institution of the focus sequence.

#### Formulae for probability calculations

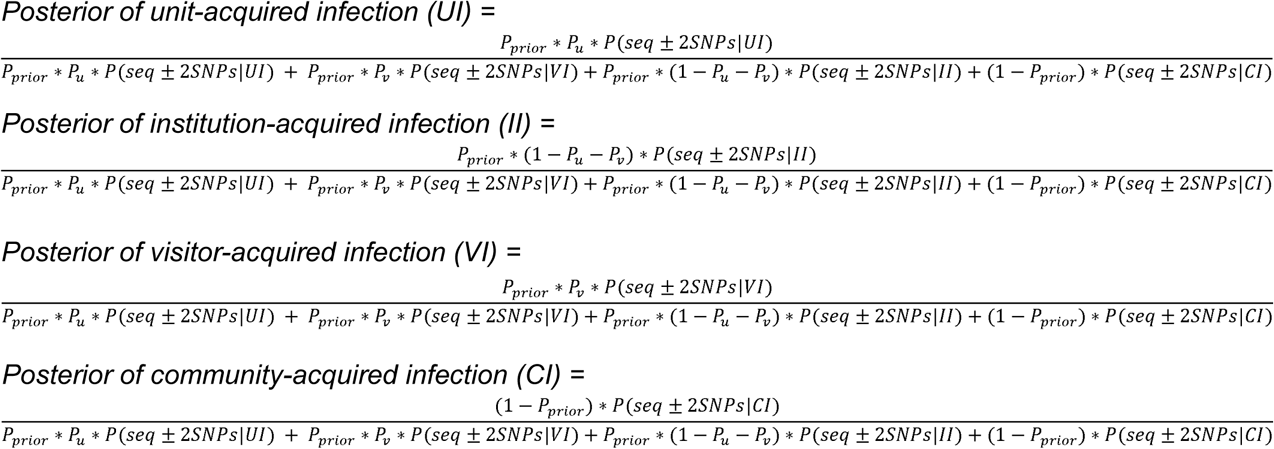

When there is a close sequence match found in any of the defined reference sets, the posterior probability estimates for UI, II, VI and CI will always sum to 1. However, when there is no close sequence match in any of the reference sets the posterior probability calculations are not valid and the algorithm will return the prior probabilities for each potential source/location of infection.

#### Further details regarding sequence matching process

The ±2 SNP threshold for a close sequence match was initially based on reports of healthcare-associated outbreak events for which this was the maximum pairwise difference within clusters (Meredith: DOI:10.1101/2020.05.08.20095687 & Rockett: DOI: 10.1101/2020.04.19.048751). The outbreak events described included sequences with up to around 3 weeks between first and last samples. This SNP threshold is also supported by calculations using the overall mutation rate of SARS-CoV-2. If we take the average mutation rate of the virus to be 24 SNPs/year (Nextstrain value 24th June, https://nextstrain.org/ncov/global?l=clock), then assuming independent (Poisson distributed) mutation events, ignoring the chance of mutations occurring at the same position in the genome and using a fixed generation time of 5 days then there is an approximate:

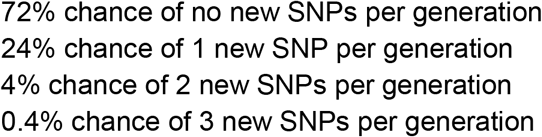

A 2 SNP threshold would therefore be expected to identify close sequence matches between direct transmission pairs in a large majority of cases. Ambiguous nucleotide positions will be considered to match if there is an overlap in the possible values for the two sequences. ‘N’ values recorded in either the focus sequence or comparison sequence will be considered to be a match at that position.

#### Further details of prior probability calculations for post-admission infection

We calculate *P*_*prior*_= *F*(*t*), where *F*() is the cumulative distribution function of a published log-normal distribution for incubation times (Lauer et al: doi:10.7326/M20-0504; μ=1.621,σ=0.418). For symptomatic HOCI cases, the IPC classifications recommended by PHE translate into the following value ranges for *P*_*prior*_:

- indeterminate HCAI: 0.11 (onset 3 days post-admission) to 0.78 (onset 7 days post-admission)
- probable HCAI: 0.86 (onset 8 days post-admission) to 0.99 (onset 14 days post-admission)
- definite HCAI: *P*_*prior*_≥0.995

For asymptomatic focus cases, we define our prior on the basis that some proportion of the cases detected will never become symptomatic (*P*_*a*_) with the remainder going on to develop symptoms within the next few days (1-*P*_*a*_). We then define our prior probability of post-admission infection in these cases as:

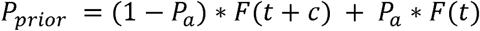

where *t* is the interval from admissionDate to sampleDate and *c* is a constant reflecting the average interval within which we expect symptoms to appear (among those cases in which they do). *P*_*a*_ is set at 0.4 based on the findings of a published review article (Oran and Topol: doi.org/10.7326/M20-3012), and *c* is set to 3 based on a combination of expert opinion of the study PIs, the known distribution of time from infection to symptom onset and expert experience of asymptomatic screening.

##### Source given post-admission infection

The model requires prior values for the probability of UI and VI given post-admission infection: *P*_*u*_ and *P*_*v*_, respectively. However, in specifying the model we define *P*_*u*_*’* as the probability of UI given post-admission infection when there are no visitors allowed on the ward, in which case the probability of VI is zero and *P*_*v*_*’*=0. If visitors are allowed on the ward for the focus case, then we set *P*_*u*_= *P*_*u*_*’*×(1-*P*_*v*_).

Based on expert opinion of the clinical co-coauthors, *P*_*u*_*’* is set to different values according to the unit/ward type of the focus sequence with single bed wards having a lower prior probability of unit post-admission infection than bay wards: 0.5 for single bed wards and 0.7 for bay wards. We assumed a *P*_*v*_ of 0.2. The *P*_*u*_ values (when visitors are allowed) are therefore: 0.4 for single bed wards and 0.56 for bay wards. The largest of the three Glasgow hospitals included comprises single-room wards, whilst the other two and the Sheffield site comprise bay wards.

#### Derivation of prior probability for post-admission infection

If we assume a uniform individual-level hazard (λ) of infection from 1st February 2020 (*t*_*0*_), whether in hospital or not, then the probability density function (PDF) of infection at time *t*_*inf*_ from this date is: λ*e*^(-λ*t*_*inf*_). The PDF of infection at time *t*_*inf*_ conditional on this occurring at any point prior to the date of symptom onset (*t*_*onset*_) is: (λ*e*^(-λ*t*_*inf*_)) / (1-*e*^(-λ*t*_*pos*_)), which is approximately 1/*t*_*onset*_ for small λ (taking the limit as λ->0). For HOCI cases, we are interested in whether t_inf_ occurred before or after the time of admission to hospital (*t*_*adm*_). Also considering the evidence provided by the known incubation time of the disease (PDF *f* and CDF *F*), we integrate over the range of possible infection dates:

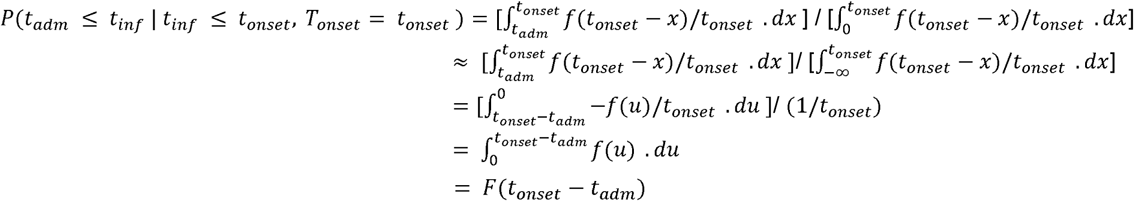

#### Geographic weighting for community reference set

##### Geographic weighting function

The weight of each sequence within the community reference set is determined by geographic distance from the residential outer postcode of the focus case, using a function of the form: weight= (1-β)*exp(-τ*communityDistanceToIndex[i]) + β, where, β takes a value between 0 and 1, and τ>0. These parameters are set based on calibration to the available community reference set at each site. The rationale for this weighting is that there is likely to be geographic clustering of viral lineages, and so newly observed community transmissions of SARS-CoV-2 are more likely to show genetic similarity to past sequences from the local area of that individual’s home than to past sequences from regions that are further away. If postcode is missing for a case in the community reference set, then distance to the focus sequence is set to 100 km.

##### Statistical model for derivation of geographic weighting parameters

The statistical model for geographic weighting is fitted separately for each study site using sequences which are strongly thought to represent community-acquired infection: all community-sampled sequences and patients presenting to A&E with COVID-19, excluding those who are recorded as being healthcare workers or who do not have an available valid outer postcode. We will refer to these sequences as the ‘calibration set’.

A statistical model is constructed to find the optimal values of β and τ to maximise the estimated probability (P_sim:i_) of a newly observed community-acquired case having a similar sequence (±2SNPs) to that observed for each sequence in the calibration set. The estimated probability in each case within the calibration set is calculated as a weighted sum of ‘close match’ indicator variables for all other sequences in the calibration set sample from 6 weeks prior up until the sample date of that case, with the weighting function defined in terms of geographic distance between residential outer postcodes and the β and τ parameters as described for the community reference set.

An overall log-likelihood function is defined using a Bernoulli distribution for each of the *n* sequences within the calibration set:

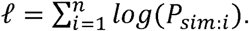

The values of β and τ that maximise ℓ were obtained for each of the study sites using the ‘bbmle’ package for R, with logit-parameterisation of β and log-parameterisation of τ.

We assume that the probability of a sequence match conditional on infection from visitor on unit/ward can be calculated using the same weighting scheme as for the probability of a sequence match conditional on community-acquired infection (i.e. P(seq±2 SNPs|CI)==P(seq±2 SNPs|VI)).

#### Additional matching on ward location history

There is the potential for the algorithm described to return large numbers of close sequences matches with the hospital as a whole, which may make it difficult for IPC teams to use the output to direct their investigations when there are no potential sources of infection identified on the same ward as the focus case. We propose a location matching procedure in order to highlight the most relevant sequence matches for further investigation. This process does not currently form part of the statistical model, meaning that it can be treated as optional functionality for the SRT in the COG-UK HOCI study, and we have restricted the input data to a simplified format in order to minimise data management requirements.

For each inpatient sample in the input meta-data for the algorithm, we specify a single string variable comprising the concatenated names of any ward locations in the ≤14 days prior to the sample date and a separate string variable with any ward locations in the ≤14 days after the sample date. For each focus case submitted to the algorithm, output is flagged if there is any match identified between the wards listed in each of these fields or the ward at time of sampling for a close sequence match in comparison to the prior and current ward locations for the focus sequence (excluding those cases were there is already matching ward location at time of sampling for each).

#### Details of phylogenetic methods

Phylogenies were produced by the grapevine pipeline (https://github.com/COG-UK/grapevine) as part of the COG-UK Consortium (https://www.cogconsortium.uk). Briefly, sequences from GISAID and those produced as part of the COG-UK Consortium are independently quality controlled and aligned to the Wuhan reference using minimap2 (https://doi.org/10.1093/bioinformatics/bty191). The two alignments are then combined, the homoplasy at site 11083 is masked and the tree is reconstructed using FastTreeMP (http://www.microbesonline.org/fasttree/). For each of the hospitals of interest, the tree is pruned to keep sequences from Scotland or Yorkshire (as relevant) and by date excluding sequences subsequent to the last “focus” patient sample date on the ward.

#### Details of SRT report format

The SRT system for prospective use needs to provide useful and appropriate feedback in both low incidence and high incidence settings for new HOCI cases. This is planned through the generation of a concise one-page PDF summary report for each focus sequence. This summary report will contain key focus sequence meta-data, information regarding the estimated probabilities for infection source and details of up to ten close sequence matches identified within the same unit/ward and/or elsewhere in the hospital.

#### Probability summary categories

The sequence matching and probability score algorithm generates probability estimates for the source of infection for the focus patient being from the current unit/ward, from elsewhere in the hospital, from the community (pre-admission) or from a visitor. These probability estimates always sum to 1. In the summary report, probability estimates for each source of infection are categorised using the following levels:

- 0-30%: low
- 30-50%: moderately low
- 50-70%: probable
- 70-85%: high
- 85-100%: very high

For clarity of presentation and communication, probability categories will not always be displayed in the summary report for all four potential sources of infection (i.e. ward/unit, elsewhere in hospital, visitor, or community). Special handling rules for specific situations are described below.

#### Close sequence matches within the same unit and/or hospital

The maximum number of close sequence matches that can be listed on the one-page summary report is 10 (for the combined sum of unit-level and institution-level matches). If the number of ward-level matches is n>5 and the total number of close sequence matches is N>10, then the number of ward-level matches is truncated at 5+max((5-(N-n)),0). If there are over ten close sequence matches in total, then the following message is displayed “Over 10 close matches; see detailed report for further information”.

Within the each the sets of unit-level and institution-level close sequence matches, ordering and priority for inclusion within the available slots is determined by the following set of criteria (in decreasing order of importance):

1. Number of SNPs relative to Wuhan strain present in comparison sequence but absent in focus sequence (fewer = higher priority)
2. Number of SNPs relative to Wuhan strain present in focus sequence but absent in comparison sequence (fewer = higher priority)
3. Whether comparison sequence is from a HCW (HCWs listed first)
4. HCAI status of comparison sequence (priority order: definite, probable, indeterminate, otherwise)
5. Samples from the past before samples in future
6. Samples from within the two weeks prior to focus sequence sample date before others
7. Number of units overlapping with focus sample’s units

#### Report messages for specific output combinations

##### No close sequence matches on unit/ward

If there are no close sequence matches to the focus sequence on their current unit/ward, then no probability category is reported for this potential infection source (the algorithm returns a zero probability in such cases, which could be misleading given uncertainty over screening and sequencing coverage). The message “No matches from within unit” is displayed. The probability score category for infection from elsewhere in the hospital is provided in such cases.

##### No close sequence matches elsewhere in hospital

If there are no close sequence matches to the focus sequence elsewhere in the hospital, then no probability category is reported for this potential infection source. The message “No matches elsewhere in hospital” is displayed.

##### No evidence of transmission within unit or hospital for probable or definite HCAI

If the estimated probability of community-acquired infection from the algorithm is >50%, but the interval from admission to symptom onset (if recorded) or sample date is ≥8 days, then the following message is displayed in place of the estimated probability of community-acquired infection “This is a probable/definite HCAI based on admission date, but we have not found genetic evidence of transmission within the hospital”.

##### Probable unit- or hospital-acquired infection with source unclear

If the posterior probability of unit-acquired infection and the posterior probability of infection from a source elsewhere in the hospital are each estimated to be <50%, but the sum of these two posterior probabilities is ≥50%, then the following message is displayed “Overall, this is a probable unit- or institution-acquired infection with source unclear”.

#### Timeline graph

The timeline graph provides a visual representation of available sequences from the same unit/ward and the same institution/hospital as the focus sequence in the period from 3 weeks prior to their sample date to 1 week after. The key indicates which sequences are close matches to the focus sequence, and the numbering corresponds to that in the tabular summary of most relevant close sequence matches.

### Results

#### Sequencing coverage in Glasgow dataset

**Figure S1.**
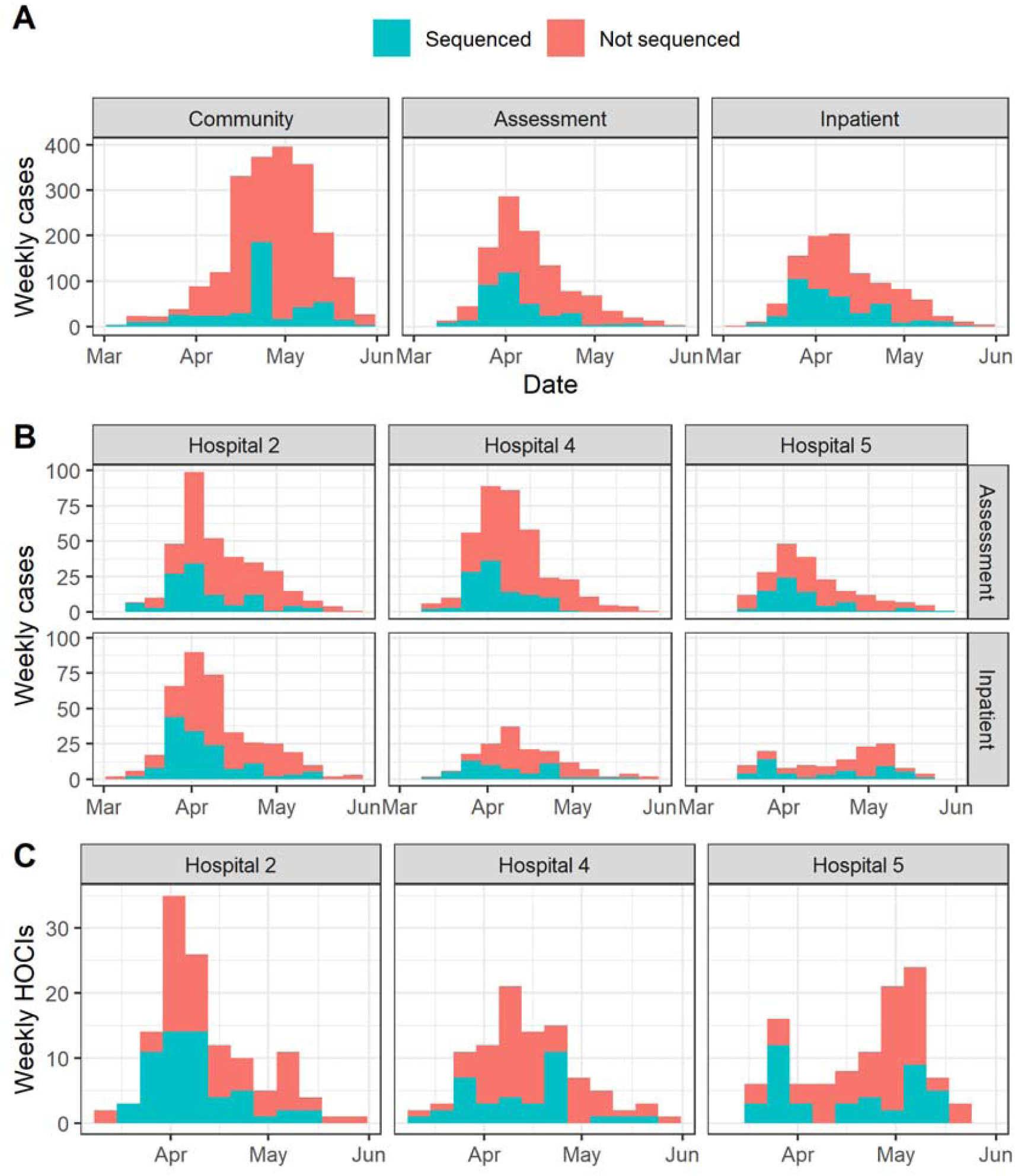
Proportion of cases sequenced in Greater Glasgow and Clyde Health Board between 1 March and 27^th^ May (with sequence available as of 23 June 2020) by location of test (A). Also displayed are the proportion of sequenced cases in the three focus hospitals subdivided by assessment and inpatient locations (B), and the proportion of HOCI cases sequenced at these hospitals (C).

#### Home residence locations and geographic model parameters

**Figure S2.**
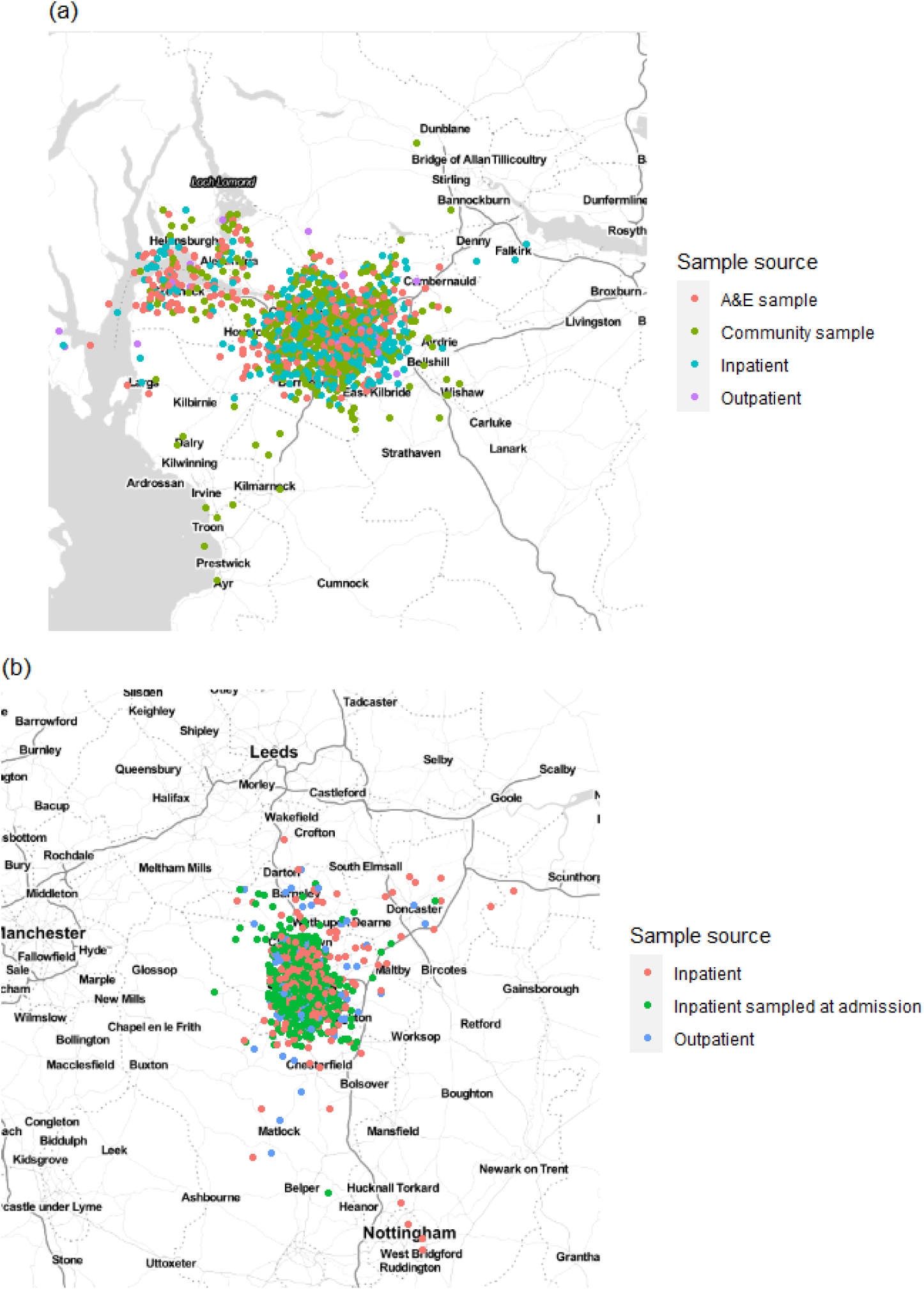
Home residence location of individuals in (a) the Glasgow dataset and (b) the Sheffield dataset, displayed by sample source (not including HCWs). Locations are analysed using only the outer postcode, and as such random jitter (within longitude and latitude of 0.05) has been added to allow display without overlap of points. Plot created using ggmap for R with map obtained from Stamen maps. For Glasgow 766 cases were included in the calibration set with estimates of τ=0.15 and β=0.0 for the geographic clustering model, whilst for Sheffield 446 cases were included in the calibration set with resulting estimates of τ=0.84 and β=0.16.

#### SNP distance distributions

For the Glasgow sequence dataset as a whole the median pairwise SNP difference among all sequences was 9, and there were 1.3%, 3.4%, 6.4% and 10.1% of pairwise comparisons with 0, ≤1, ≤2 and ≤3 SNP differences, respectively. For the Sheffield dataset as a whole the median pairwise SNP difference among all sequences was 8, and there were 1.2%, 3.3%, 6.5% and 10.8% of pairwise comparisons with 0, ≤1, ≤2 and ≤3 SNP differences, respectively.

**Figure S3.**
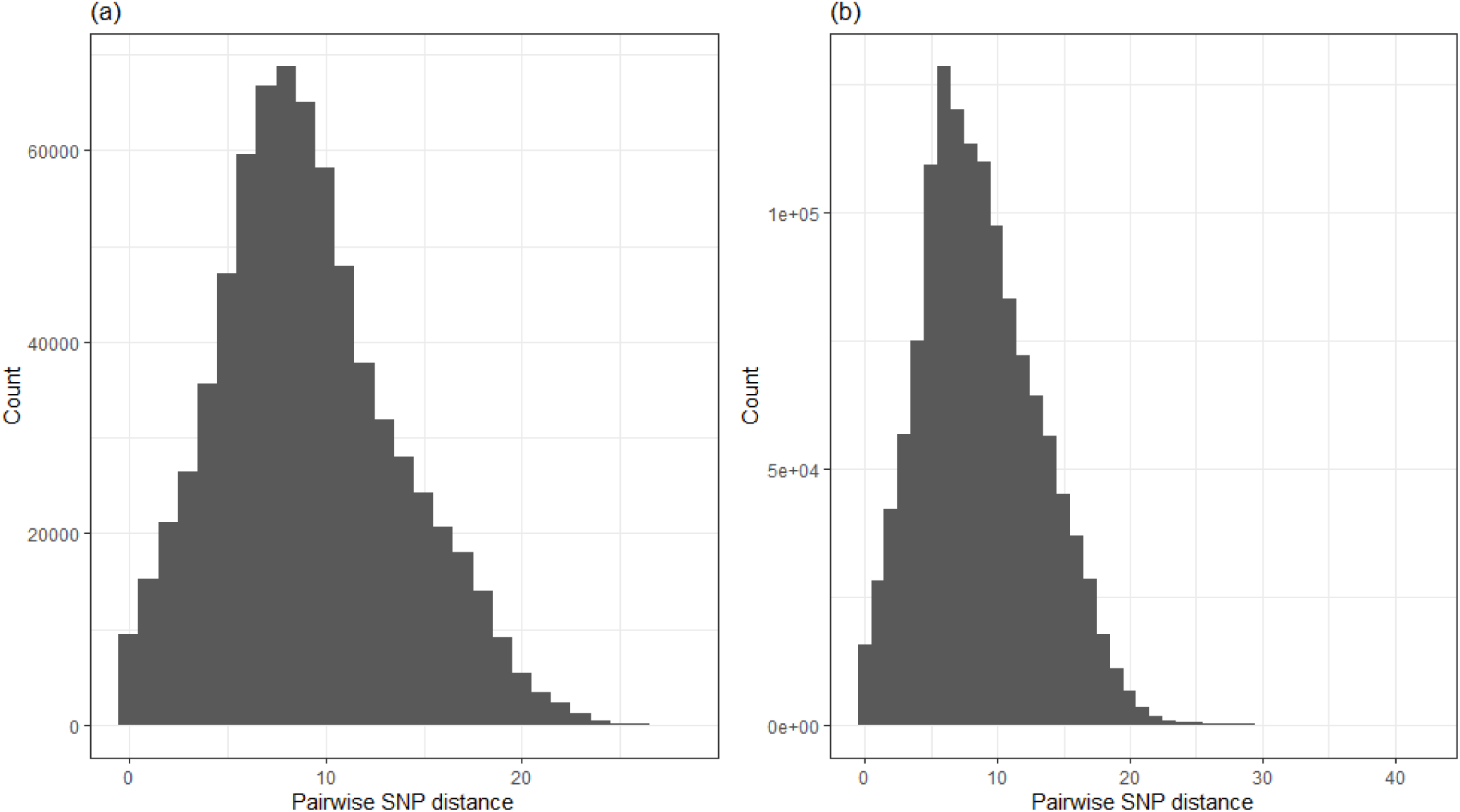
Frequency plot of all pairwise SNP differences among (a) all 1199 sequences in the Glasgow dataset and (b) all 1629 analysed sequences in the Sheffield dataset.

#### Examples of SRT reports

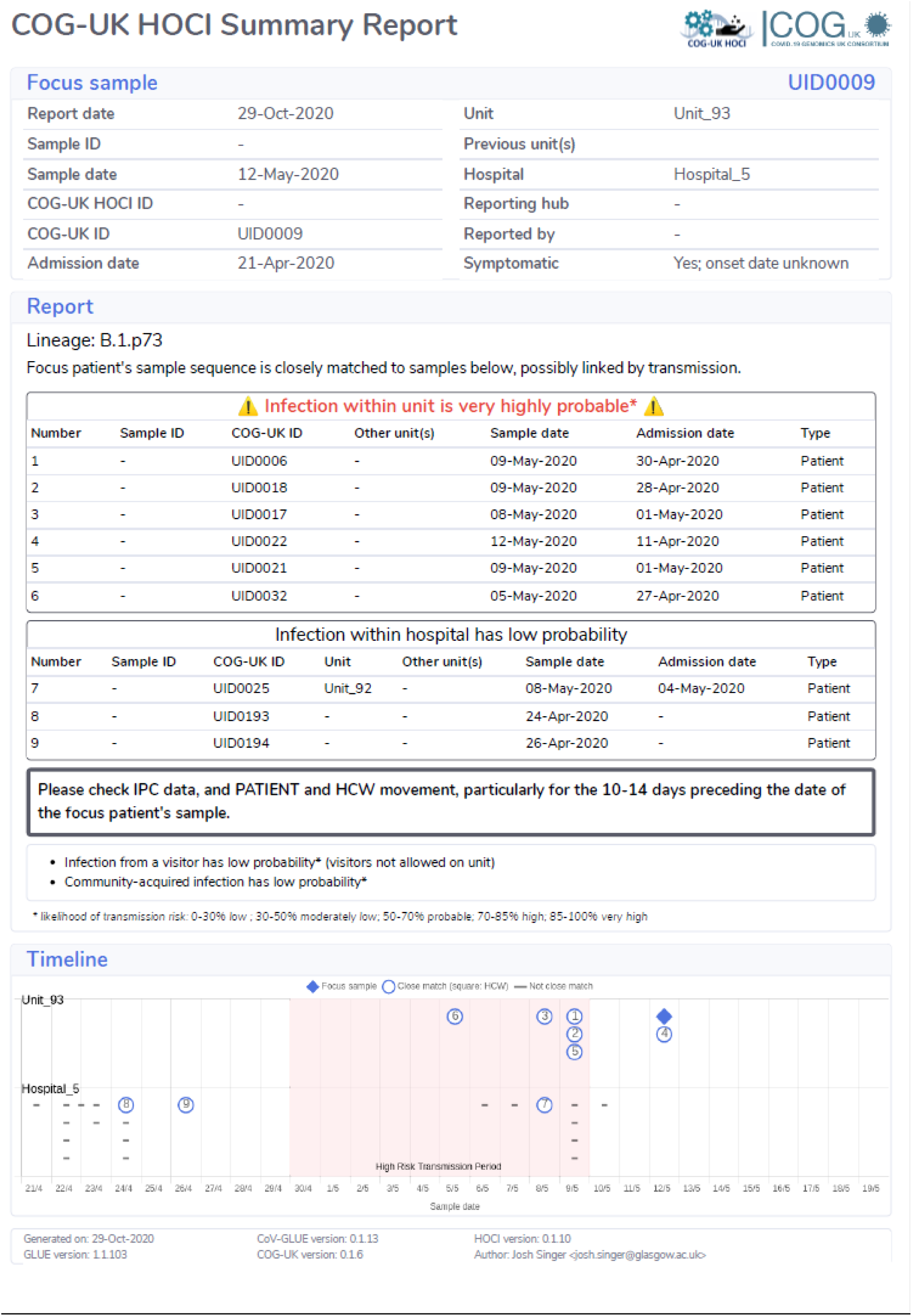
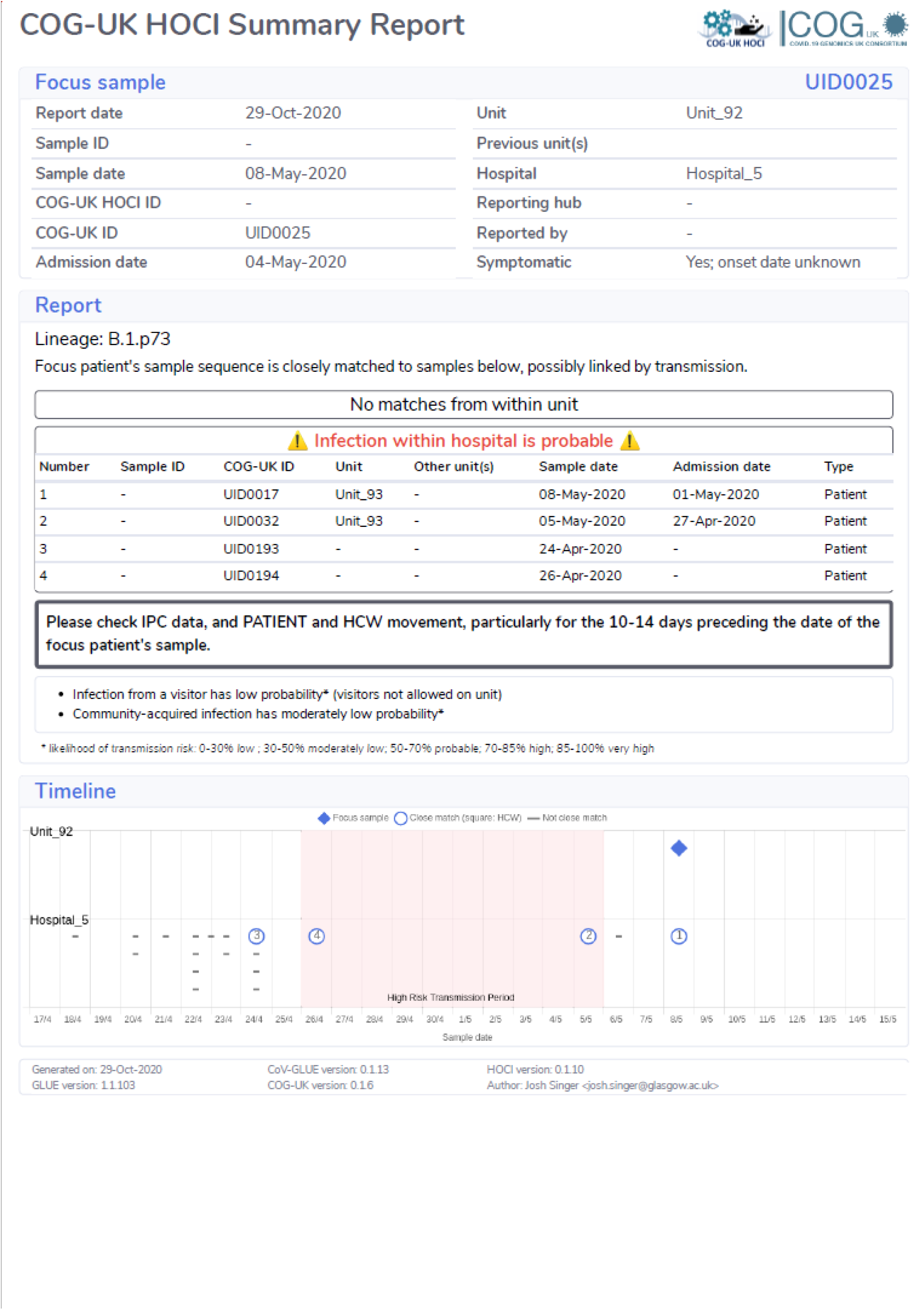

#### The COVID-19 Genomics UK (COG-UK) consortium

**Funding acquisition, leadership, supervision, metadata curation, project administration, samples, logistics, Sequencing, analysis, and Software and analysis tools:**

Dr Thomas R Connor PhD^33, 34^, and Professor Nicholas J Loman PhD^15^.

**Leadership, supervision, sequencing, analysis, funding acquisition, metadata curation, project administration, samples, logistics, and visualisation:**

Dr Samuel C Robson Ph.D ^68^.

**Leadership, supervision, project administration, visualisation, samples, logistics, metadata curation and software and analysis tools:**

Dr Tanya Golubchik PhD ^27^.

**Leadership, supervision, metadata curation, project administration, samples, logistics sequencing and analysis:**

Dr M. Estee Torok FRCP ^8, 10^.

**Project administration, metadata curation, samples, logistics, sequencing, analysis, and software and analysis tools:**

Dr William L Hamilton PhD ^8, 10^.

**Leadership, supervision, samples logistics, project administration, funding acquisition sequencing and analysis:**

Dr David Bonsall PhD ^27^.

**Leadership and supervision, sequencing, analysis, funding acquisition, visualisation and software and analysis tools:**

Dr Ali R Awan PhD ^74^.

**Leadership and supervision, funding acquisition, sequencing, analysis, metadata curation, samples and logistics:**

Dr Sally Corden PhD ^33^.

**Leadership supervision, sequencing analysis, samples, logistics, and metadata curation:**

Professor Ian Goodfellow PhD ^11^.

**Leadership, supervision, sequencing, analysis, samples, logistics, and Project administration:**

Professor Darren L Smith PhD ^60, 61^.

**Project administration, metadata curation, samples, logistics, sequencing and analysis:**

Dr Martin D Curran PhD ^14^, and Dr Surendra Parmar PhD ^14^.

**Samples, logistics, metadata curation, project administration sequencing and analysis:**

Dr James G Shepherd MBChB MRCP ^21^.

**Sequencing, analysis, project administration, metadata curation and software and analysis tools:**

Dr Matthew D Parker PhD ^38^ and Dr Dinesh Aggarwal MRCP ^1, 2, 3^.

**Leadership, supervision, funding acquisition, samples, logistics, and metadata curation:**

Dr Catherine Moore^33^.

**Leadership, supervision, metadata curation, samples, logistics, sequencing and analysis:**

Dr Derek J Fairley PhD ^6, 88^, Professor Matthew W Loose PhD ^54^, and Joanne Watkins MSc ^33^.

**Metadata curation, sequencing, analysis, leadership, supervision and software and analysis tools:**

Dr Matthew Bull PhD^33^, and Dr Sam Nicholls PhD ^15^.

**Leadership, supervision, visualisation, sequencing, analysis and software and analysis tools:**

Professor David M Aanensen PhD ^1, 30^.

**Sequencing, analysis, samples, logistics, metadata curation, and visualisation:**

Dr Sharon Glaysher ^70^.

**Metadata curation, sequencing, analysis, visualisation, software and analysis tools:**

Dr Matthew Bashton PhD ^60^, and Dr Nicole Pacchiarini PhD ^33^.

**Sequencing, analysis, visualisation, metadata curation, and software and analysis tools**: Dr Anthony P Underwood PhD ^1, 30^.

**Funding acquisition, leadership, supervision and project administration:**

Dr Thushan I de Silva PhD ^38^, and Dr Dennis Wang PhD ^38^.

**Project administration, samples, logistics, leadership and supervision**:

Dr Monique Andersson PhD ^28^, Professor Anoop J Chauhan ^70^, Dr Mariateresa de Cesare PhD^26^, Dr Catherine Ludden ^1,3^, and Dr Tabitha W Mahungu FRCPath ^91^.

**Sequencing, analysis, project administration and metadata curation:**

Dr Rebecca Dewar PhD ^20^, and Martin P McHugh MSc^20^.

**Samples, logistics, metadata curation and project administration:**

Dr Natasha G Jesudason MBChB MRCP FRCPath ^21^, Dr Kathy K Li MBBCh FRCPath ^21^, Dr Rajiv N Shah BMBS MRCP MSc ^21^, and Dr Yusri Taha MD, PhD ^66^.

**Leadership, supervision, funding acquisition and metadata curation:**

Dr Kate E Templeton PhD ^20^.

**Leadership, supervision, funding acquisition, sequencing and analysis:**

Dr Simon Cottrell PhD ^33^, Dr Justin O’Grady PhD ^51^, Professor Andrew Rambaut DPhil ^19^, and Professor Colin P Smith PhD^93^.

**Leadership, supervision, metadata curation**, **sequencing and analysis:**

Professor Matthew T.G. Holden PhD ^87^, and Professor Emma C Thomson PhD/FRCP ^21^.

**Leadership, supervision, samples, logistics and metadata curation**: Dr Samuel Moses MD ^81, 82^.

**Sequencing, analysis, leadership, supervision, samples and logistics:**

Dr Meera Chand ^7^, Dr Chrystala Constantinidou PhD ^71^, Professor Alistair C Darby PhD ^46^, Professor Julian A Hiscox PhD ^46^, Professor Steve Paterson PhD ^46^, and Dr Meera Unnikrishnan PhD ^71^.

**Sequencing, analysis, leadership and supervision and software and analysis tools:**

Dr Andrew J Page PhD ^51^, and Dr Erik M Volz PhD^96^.

**Samples, logistics, sequencing, analysis and metadata curation:**

Dr Charlotte J Houldcroft PhD ^8^, Dr Aminu S Jahun PhD ^11^, Dr James P McKenna PhD ^88^, Dr Luke W Meredith PhD ^11^, Dr Andrew Nelson PhD ^61^, Sarojini Pandey MSc ^72^, and Dr Gregory R Young PhD ^60^.

**Sequencing, analysis, metadata curation, and software and analysis tools:**

Dr Anna Price PhD^34^, Dr Sara Rey PhD ^33^, Dr Sunando Roy PhD ^41^, Dr Ben Temperton Ph.D ^49^, and Matthew Wyles ^38^.

**Sequencing, analysis, metadata curation and visualisation:**

Stefan Rooke MSc^19^, and Dr Sharif Shaaban PhD^87^.

**Visualisation, sequencing, analysis and software and analysis tools:**

Dr Helen Adams PhD ^35^, Dr Yann Bourgeois Ph.D ^69^, Dr Katie F Loveson Ph.D ^68^, Áine O’Toole MSc^19^, and Richard Stark MSc ^71^.

**Project administration, leadership and supervision:**

Dr Ewan M Harrison PhD ^1, 3^, David Heyburn ^33^, and Professor Sharon J Peacock ^2, 3^

**Project administration and funding acquisition:**

Dr David Buck PhD^26^, and Michaela John BSc Hons ^36^

**Sequencing, analysis and project administration:**

Dorota Jamrozy ^1^, and Dr Joshua Quick PhD ^15^

**Samples, logistics, and project administration:**

Dr Rahul Batra MD^78^, Katherine L Bellis BSc (Hons) ^1, 3^, Beth Blane BSc ^3^, Sophia T Girgis MSc ^3^, Dr Angie Green PhD ^26^, Anita Justice MSc ^28^, Dr Mark Kristiansen PhD ^41^, and Dr Rachel J Williams PhD ^41^.

**Project administration, software and analysis tools:**

Radoslaw Poplawski BSc ^15^.

**Project administration and visualisation:**

Dr Garry P Scarlett Ph.D^69^.

**Leadership, supervision, and funding acquisition:**

Professor John A Todd PhD ^26^, Dr Christophe Fraser PhD ^27^,Professor Judith Breuer MD ^40,41^, Professor Sergi Castellano PhD ^41^, Dr Stephen L Michell PhD ^49^, Professor Dimitris Gramatopoulos PhD, FRCPath^73^, and Dr Jonathan Edgeworth PhD, FRCPath ^78^.

**Leadership, supervision and metadata curation:**

Dr Gemma L Kay PhD ^51^.

**Leadership, supervision, sequencing and analysis:**

Dr Ana da Silva Filipe PhD ^21^, Dr Aaron R Jeffries PhD ^49^, Dr Sascha Ott PhD ^71^, Professor Oliver Pybus ^24^, Professor David L Robertson PhD ^21^, Dr David A Simpson PhD ^6^, and Dr Chris Williams MB BS^33^.

**Samples, logistics, leadership and supervision:**

Dr Cressida Auckland FRCPath ^50^, Dr John Boyes MBChB^83^, Dr Samir Dervisevic FRCPath^52^, Professor Sian Ellard FRCPath^49, 50^, Dr Sonia Goncalves^1^, Dr Emma J Meader FRCPath ^51^, Dr Peter Muir PhD^2^, Dr Husam Osman PhD ^95^, Reenesh Prakash MPH^52^, Dr Venkat Sivaprakasam PhD^18^, and Dr Ian B Vipond PhD^2^.

**Leadership, supervision and visualisation**

Dr Jane AH Masoli MBChB ^49, 50^.

**Sequencing, analysis and metadata curation**

Dr Nabil-Fareed Alikhan PhD ^51^, Matthew Carlile BSc ^54^, Dr Noel Craine DPhil ^33^, Dr Sam T Haldenby PhD ^46^, Dr Nadine Holmes PhD ^54^, Professor Ronan A Lyons MD ^37^, Dr Christopher Moore PhD ^54^, Malorie Perry MSc ^33^, Dr Ben Warne MRCP^80^, and Dr Thomas Williams MD ^19^.

**Samples, logistics and metadata curation:**

Dr Lisa Berry PhD ^72^, Dr Andrew Bosworth PhD ^95^, Dr Julianne Rose Brown PhD^40^, Sharon Campbell MSc^67^, Dr Anna Casey PhD ^17^, Dr Gemma Clark PhD ^56^, Jennifer Collins BSc ^66^, Dr Alison Cox PhD ^43, 44^, Thomas Davis MSc ^84^, Gary Eltringham BSc ^66^, Dr Cariad Evans ^38, 39^, Dr Clive Graham MD ^64^, Dr Fenella Halstead PhD ^18^, Dr Kathryn Ann Harris PhD ^40^, Dr Christopher Holmes PhD ^58^, Stephanie Hutchings ^2^, Professor Miren Iturriza-Gomara PhD ^46^, Dr Kate Johnson ^38, 39^, Katie Jones MSc ^72^, Dr Alexander J Keeley MRCP ^38^, Dr Bridget A Knight PhD ^49, 50^, Cherian Koshy MSc, CSci, FIBMS ^90^, Steven Liggett ^63^, Hannah Lowe MSc ^81^, Dr Anita O Lucaci PhD ^46^, Dr Jessica Lynch PhD MBChB ^25, 29^, Dr Patrick C McClure PhD ^55^, Dr Nathan Moore MBChB ^31^, Matilde Mori BSc ^25, 29, 32^, Dr David G Partridge FRCP, FRCPath ^38, 39^, Pinglawathee Madona ^43, 44^, Hannah M Pymont MSc ^2^, Dr Paul Anthony Randell MBBCh ^43, 44^, Dr Mohammad Raza ^38, 39^, Felicity Ryan MSc ^81^, Dr Robert Shaw FRCPath ^28^, Dr Tim J Sloan PhD ^57^, and Emma Swindells BSc ^65^.

**Sequencing, analysis, Samples and logistics:**

Alexander Adams BSc ^33^, Dr Hibo Asad PhD ^33^, Alec Birchley MSc ^33^, Tony Thomas Brooks BSc (Hons) ^41^, Dr Giselda Bucca PhD ^93^, Ethan Butcher ^70^, Dr Sarah L Caddy PhD ^13^, Dr Laura G Caller PhD ^2, 3, 12^, Yasmin Chaudhry BSc ^11^, Jason Coombes BSc (HONS) ^33^, Michelle Cronin ^33^, Patricia L Dyal MPhil ^41^, Johnathan M Evans MSc ^33^,Laia Fina ^33^, Bree Gatica-Wilcox MPhil ^33^, Dr Iliana Georgana PhD ^11^, Lauren Gilbert A-Levels ^33^, Lee Graham BSc ^33^, Danielle C Groves BA ^38^, Grant Hall BSc ^11^, Ember Hilvers MPH ^33^, Dr Myra Hosmillo PhD ^11^, Hannah Jones ^33^, Sophie Jones MSc ^33^, Fahad A Khokhar BSc ^13^, Sara Kumziene-Summerhayes MSc ^33^, George MacIntyre-Cockett BSc ^26^, Dr Rocio T Martinez Nunez PhD ^94^, Dr Caoimhe McKerr PhD ^33^, Dr Claire McMurray PhD ^15^, Dr Richard Myers ^7^, Yasmin Nicole Panchbhaya BSc ^41^, Malte L Pinckert MPhil ^11^, Amy Plimmer ^33^, Dr Joanne Stockton PhD ^15^, Sarah Taylor ^33^, Dr Alicia Thornton ^7^, Amy Trebes MSc ^26^, Alexander J Trotter MRes ^51^, Helena Jane Tutill BSc ^41^, Charlotte A Williams BSc ^41^, Anna Yakovleva BSc ^11^ and Dr Wen C Yew PhD ^62^.

**Sequencing, analysis and software and analysis tools:**

Dr Mohammad T Alam PhD ^71^, Dr Laura Baxter PhD ^71^, Olivia Boyd MSc ^96^, Dr Fabricia F. Nascimento PhD ^96^, Timothy M Freeman MPhil ^38^, Lily Geidelberg MSc ^96^, Dr Joseph Hughes PhD ^21^, David Jorgensen MSc ^96^, Dr Benjamin B Lindsey MRCP ^38^, Dr Richard J Orton PhD ^21^, Dr Manon Ragonnet-Cronin PhD ^96^ Joel Southgate MSc ^33, 34^, and Dr Sreenu Vattipally PhD ^21^.

**Samples, logistics and software and analysis tools:**

Dr Igor Starinskij MSc MRCP ^23^.

**Visualisation and software and analysis tools:**

Dr Joshua B Singer PhD ^21^, Dr Khalil Abudahab PhD ^1, 30^, Leonardo de Oliveira Martins PhD^51^, Dr Thanh Le-Viet PhD ^51^, Mirko Menegazzo ^30^, Ben EW Taylor Meng ^1, 30^, and Dr Corin A Yeats PhD ^30^.

**Project Administration:**

Sophie Palmer ^3^, Carol M Churcher ^3^, Dr Alisha Davies ^33^, Elen De Lacy MSc ^33^, Fatima Downing ^33^, Sue Edwards ^33^, Dr Nikki Smith PhD ^38^, Dr Francesc Coll PhD^97^, Dr Nazreen F Hadjirin PhD^3^ and Dr Frances Bolt PhD ^44, 45^.

**Leadership and supervision:**

Dr. Alex Alderton ^1^, Dr Matt Berriman ^1^, Ian G Charles ^51^, Dr Nicholas Cortes MBChB ^31^, Dr Tanya Curran PhD ^88^, Prof John Danesh ^1^, Dr Sahar Eldirdiri MBBS, MSC FRCPath ^84^, Dr Ngozi Elumogo FRCPath ^52^, Prof Andrew Hattersley FRS ^49, 50^, Professor Alison Holmes MD ^44, 45^, Dr Robin Howe ^33^, Dr Rachel Jones ^33^, Anita Kenyon MSc ^84^, Prof Robert A Kingsley PhD ^51^, Professor Dominic Kwiatkowski ^1, 9^, Dr Cordelia Langford^1^, Dr Jenifer Mason MBBS ^48^, Dr Alison E Mather PhD ^51^, Lizzie Meadows MA ^51^, Dr Sian Morgan FRCPath ^36^, Dr James Price PhD ^44, 45^, Trevor I Robinson MSc ^48^, Dr Giri Shankar ^33^, John Wain ^51^, and Dr Mark A Webber PhD^51^.

**Metadata curation:**

Dr Declan T Bradley PhD ^5, 6^, Dr Michael R Chapman PhD ^1, 3, 4^, Dr Derrick Crooke ^28^, Dr David Eyre PhD ^28^, Professor Martyn Guest PhD^34^, Huw Gulliver ^34^, Dr Sarah Hoosdally ^28^, Dr Christine Kitchen PhD ^34^, Dr Ian Merrick PhD ^34^, Siddharth Mookerjee MPH ^44, 45^, Robert Munn BSc ^34^, Professor Timothy Peto PhD^28^, Will Potter ^52^, Dr Dheeraj K Sethi MBBS ^52^, Wendy Smith ^56^, Dr Luke B Snell MB BS ^75, 94^, Dr Rachael Stanley PhD ^52^, Claire Stuart ^52^ and Dr Elizabeth Wastenge MD ^20^.

**Sequencing and analysis:**

Dr Erwan Acheson PhD ^6^, Safiah Afifi BSc ^36^, Dr Elias Allara MD PhD ^2, 3^, Dr Roberto Amato ^1^, Dr Adrienn Angyal PhD^38^, Dr Elihu Aranday-Cortes PhD/DVM^21^, Cristina Ariani ^1^, Jordan Ashworth ^19^, Dr Stephen Attwood ^24^, Alp Aydin MSci ^51^, David J Baker BEng ^51^, Dr Carlos E Balcazar PhD ^19^, Angela Beckett MSc ^68^ Robert Beer BSc ^36^, Dr Gilberto Betancor PhD^76^, Emma Betteridge ^1^, Dr David Bibby ^7^, Dr Daniel Bradshaw ^7^, Catherine Bresner Bsc(Hons) ^34^, Dr Hannah E Bridgewater PhD ^71^, Alice Broos BSc (Hons) ^21^, Dr Rebecca Brown PhD ^38^, Dr Paul E Brown PhD ^71^, Dr Kirstyn Brunker PhD ^22^, Dr Stephen N Carmichael PhD ^21^, Jeffrey K. J. Cheng MSc ^71^, Dr Rachel Colquhoun DPhil ^19^, Dr Gavin Dabrera ^7^, Dr Johnny Debebe PhD ^54^, Eleanor Drury ^1^, Dr Louis du Plessis ^24^, Richard Eccles MSc ^46^, Dr Nicholas Ellaby ^7^, Audrey Farbos MSc ^49^, Ben Farr ^1^, Dr Jacqueline Findlay PhD ^41^, Chloe L Fisher MSc ^74^, Leysa Marie Forrest MSc ^41^, Dr Sarah Francois ^24^, Lucy R. Frost BSc ^71^, William Fuller BSc ^34^, Dr Eileen Gallagher ^7^, Dr Michael D Gallagher PhD^19^, Matthew Gemmell MSc ^46^, Dr Rachel AJ Gilroy PhD ^51^, Scott Goodwin ^1^, Dr Luke R Green PhD ^38^, Dr Richard Gregory PhD ^46^, Dr Natalie Groves ^7^, Dr James W Harrison PhD ^49^, Hassan Hartman ^7^, Dr Andrew R Hesketh PhD^93^,Verity Hill ^19^, Dr Jonathan Hubb ^7^, Dr Margaret Hughes PhD^46^, Dr David K Jackson ^1^, Dr Ben Jackson PhD ^19^, Dr Keith James ^1^, Natasha Johnson BSc (Hons)^21^,Ian Johnston ^1^, Jon-Paul Keatley ^1^, Dr Moritz Kraemer ^24^, Dr Angie Lackenby ^7^, Dr Mara Lawniczak ^1^, Dr David Lee ^7^, Rich Livett ^1^, Stephanie Lo ^1^, Daniel Mair BSc (Hons) ^21^, Joshua Maksimovic FD sport science ^36^, Nikos Manesis ^7^, Dr Robin Manley Ph.D ^49^, Dr Carmen Manso ^7^,Dr Angela Marchbank BSc ^34^, Dr Inigo Martincorena ^1^, Dr Tamyo Mbisa ^7^, Kathryn McCluggage MSC ^36^,Dr JT McCrone PhD ^19^, Shahjahan Miah ^7^, Michelle L Michelsen BSc ^49^, Dr Mari Morgan PhD ^33^, Dr Gaia Nebbia PhD, FRCPath ^78^,Charlotte Nelson MSc ^46^, Jenna Nichols BSc (Hons) ^21^, Dr Paola Niola PhD ^41^, Dr Kyriaki Nomikou PhD^21^, Steve Palmer ^1^, Dr. Naomi Park ^1^, Dr Yasmin A Parr PhD^21^, Dr Paul J Parsons PhD ^38^, Vineet Patel ^7^, Dr. Minal Patel ^1^, Clare Pearson MSc ^2, 1^, Dr Steven Platt ^7^, Christoph Puethe ^1^, Dr. Mike Quail ^1^,Dr JaynaRaghwani ^24^, Dr Lucille Rainbow PhD ^46^, Shavanthi Rajatileka ^1^, Dr Mary Ramsay ^7^, Dr Paola C Resende Silva PhD ^41, 42^, Steven Rudder 51, Dr Chris Ruis ^3^, Dr Christine M Sambles PhD ^49^, Dr Fei Sang PhD ^54^, Dr Ulf Schaefer^7^, Dr Emily Scher PhD ^19^, Dr. Carol Scott ^1^, Lesley Shirley ^1^, Adrian W Signell BSc ^76^, John Sillitoe ^1^, Christen Smith ^1^, Dr Katherine L Smollett PhD ^21^, Karla Spellman FD ^36^, Thomas D Stanton BSc ^19^, Dr David J Studholme PhD ^49^, Ms Grace Taylor-Joyce BSc ^71^, Dr Ana P Tedim PhD ^51^, Dr Thomas Thompson PhD^6^, Dr Nicholas M Thomson PhD ^51^, Scott Thurston^1^, Lily Tong PhD ^21^, Gerry Tonkin-Hill ^1^, Rachel M Tucker MSc ^38^, Dr Edith E Vamos PhD ^4^,Dr Tetyana Vasylyeva^24^, Joanna Warwick-Dugdale BSc ^49^, Danni Weldon ^1^, Dr Mark Whitehead PhD ^46^, Dr David Williams ^7^,Dr Kathleen A Williamson PhD^19^,Harry D Wilson BSc ^76^,Trudy Workman HNC ^34^, Dr Muhammad Yasir PhD ^51^, Dr Xiaoyu Yu PhD ^19^, and Dr Alex Zarebski ^24^.

**Samples and logistics:**

Dr Evelien M Adriaenssens PhD ^51^, Dr Shazaad S Y Ahmad MSc ^2, 47^, Adela Alcolea-Medina MPharm ^59, 77^, Dr John Allan PhD^60^, Dr Patawee Asamaphan PhD^21^, Laura Atkinson MSc ^40^, Paul Baker MD ^63^, Professor Jonathan Ball PhD ^55^, Dr Edward Barton MD^64^, Dr. Mathew A Beale^1^, Dr. Charlotte Beaver^1^, Dr Andrew Beggs PhD^16^, Dr Andrew Bell PhD^51^, Duncan J Berger ^1^, Dr Louise Berry. ^56^, Claire M Bewshea MSc ^49^, Kelly Bicknell ^70^, Paul Bird ^58^, Dr Chloe Bishop ^7^, Dr Tim Boswell ^56^, Cassie Breen BSc^48^, Dr Sarah K Buddenborg^1^, Dr Shirelle Burton-Fanning MD ^66,^ Dr Vicki Chalker ^7^, Dr Joseph G Chappell PhD ^55^, Themoula Charalampous MSc ^78, 94^, Claire Cormie^3^, Dr Nick Cortes PhD^29, 25^, Dr Lindsay J Coupland PhD ^52^, Angela Cowell MSc^48^, Dr Rose K Davidson PhD ^53^, Joana Dias MSc^3^, Dr Maria Diaz PhD^51^, Thomas Dibling^1^, Matthew J Dorman^1^, Dr Nichola Duckworth^57^, Scott Elliott^70^, Sarah Essex^63^, Karlie Fallon ^58^, Theresa Feltwell ^8^, Dr Vicki M Fleming PhD ^56^, Sally Forrest BSc ^3^, Luke Foulser^1^, Maria V Garcia-Casado^1^, Dr Artemis Gavriil PhD ^41^, Dr Ryan P George PhD^47^, Laura Gifford MSc ^33^, Harmeet K Gill PhD^3^, Jane Greenaway MSc^65^, Luke Griffith Bsc^53^, Ana Victoria Gutierrez^51^, Dr Antony D Hale MBBS^85^, Dr Tanzina Haque FRCPath, PhD^91^, Katherine L Harper MBiol^85^, Dr Ian Harrison ^7^, Dr Judith Heaney PhD^89^, Thomas Helmer ^58^, Ellen E Higginson PhD ^3^, Richard Hopes ^2^, Dr Hannah C Howson-Wells PhD ^56^, Dr Adam D Hunter ^1^, Robert Impey ^70^, Dr Dianne Irish-Tavares FRCPath ^91^, David A Jackson^1^, Kathryn A Jackson MSc ^46^, Dr Amelia Joseph ^56^, Leanne Kane ^1^, Sally Kay ^1^, Leanne M Kermack MSc ^3^, Manjinder Khakh ^56^, Dr Stephen P Kidd PhD^29, 25,31^,, Dr Anastasia Kolyva PhD ^51^, Jack CD Lee BSc ^40^, Laura Letchford ^1^, Nick Levene MSc^79^, Dr LisaJ Levett PhD ^89^, Dr Michelle M Lister PhD ^56^, Allyson Lloyd ^70^, Dr Joshua Loh PhD^60^, Dr Louissa R Macfarlane-Smith PhD^85^, Dr Nicholas W Machin MSc ^2, 47^, Mailis Maes M.phil^3^, Dr Samantha McGuigan ^1^, Liz McMinn ^1^, Dr Lamia Mestek-Boukhibar D.Phil ^41^, Dr Zoltan Molnar PhD ^6^, Lynn Monaghan ^79^, Dr Catrin Moore ^27^, Plamena Naydenova BSc ^3^, Alexandra S Neaverson ^1^, Dr. Rachel Nelson PhD ^1^, Marc O Niebel MSc^21^, Elaine O’Toole BSc ^48^, Debra Padgett BSc ^64^, Gaurang Patel ^1^, Dr Brendan AI Payne MD ^66^, Liam Prestwood ^1^, Dr Veena Raviprakash MD^67^, Nicola Reynolds PhD^86^ Dr Alex Richter PhD ^16^, Dr Esther Robinson PhD^95^, Dr Hazel A Rogers^1^, Dr Aileen Rowan PhD ^96^, Garren Scott BSc ^64^, Dr Divya Shah PhD^40^, Nicola Sheriff BSc ^67^, Dr Graciela Sluga MD - MSc^92^, Emily Souster^1^, Dr. Michael Spencer-Chapman^1^, Sushmita Sridhar BSc^1, 3^, Tracey Swingler ^53^, Dr Julian Tang^58^, Professor Graham P Taylor DSc^96^, Dr Theocharis Tsoleridis PhD^55^, Dr Lance Turtle PhD MRCP^46^, Dr Sarah Walsh ^57^, Dr Michelle Wantoch PhD ^86^, Joanne Watts BSc^48^, Dr Sheila Waugh MD^66^, Sam Weeks^41^, Dr Rebecca Williams BMBS ^31^, Dr Iona Willingham^56^, Dr Emma L Wise PhD ^25, 29, 31^, Victoria Wright BSc ^54^, Dr Sarah Wyllie ^70^, and Jamie Young BSc ^3^.

**Software and analysis tools**

Amy Gaskin MSc^33^, Dr Will Rowe PhD ^15^, and Dr Igor Siveroni PhD^96^.

**Visualisation:**

Dr Robert Johnson PhD ^96^.

**1** Wellcome Sanger Institute, **2** Public Health England, **3** University of Cambridge, **4** Health Data Research UK, Cambridge, **5** Public Health Agency, Northern Ireland, **6** Queen’s University Belfast **7** Public Health England Colindale, **8** Department of Medicine, University of Cambridge, **9** University of Oxford, **10** Departments of Infectious Diseases and Microbiology, Cambridge University Hospitals NHS Foundation Trust; Cambridge, UK, **11** Division of Virology, Department of Pathology, University of Cambridge, **12** The Francis Crick Institute, **13** Cambridge Institute for Therapeutic Immunology and Infectious Disease, Department of Medicine, **14** Public Health England, Clinical Microbiology and Public Health Laboratory, Cambridge, UK, **15** Institute of Microbiology and Infection, University of Birmingham, **16** University of Birmingham, **17** Queen Elizabeth Hospital, **18** Heartlands Hospital, **19** University of Edinburgh, **20** NHS Lothian, **21** MRC-University of Glasgow Centre for Virus Research, **22** Institute of Biodiversity, Animal Health & Comparative Medicine, University of Glasgow, **23** West of Scotland Specialist Virology Centre, **24** Dept Zoology, University of Oxford, **25** University of Surrey, **26** Wellcome Centre for Human Genetics, Nuffield Department of Medicine, University of Oxford, **27** Big Data Institute, Nuffield Department of Medicine, University of Oxford, **28** Oxford University Hospitals NHS Foundation Trust, **29** Basingstoke Hospital, **30** Centre for Genomic Pathogen Surveillance, University of Oxford, **31** Hampshire Hospitals NHS Foundation Trust, **32** University of Southampton, **33** Public Health Wales NHS Trust, **34** Cardiff University, **35** Betsi Cadwaladr University Health Board, **36** Cardiff and Vale University Health Board, **37** Swansea University, **38** University of Sheffield, **39** Sheffield Teaching Hospitals, **40** Great Ormond Street NHS Foundation Trust, **41** University College London, **42** Oswaldo Cruz Institute, Rio de Janeiro **43** North West London Pathology, **44** Imperial College Healthcare NHS Trust, **45** NIHR Health Protection Research Unit in HCAI and AMR, Imperial College London, **46** University of Liverpool, **47** Manchester University NHS Foundation Trust, **48** Liverpool Clinical Laboratories, **49** University of Exeter, **50** Royal Devon and Exeter NHS Foundation Trust, **51** Quadram Institute Bioscience, University of East Anglia, **52** Norfolk and Norwich University Hospital, **53** University of East Anglia, **54** Deep Seq, School of Life Sciences, Queens Medical Centre, University of Nottingham, **55** Virology, School of Life Sciences, Queens Medical Centre, University of Nottingham, **56** Clinical Microbiology Department, Queens Medical Centre, **57** PathLinks, Northern Lincolnshire & Goole NHS Foundation Trust, **58** Clinical Microbiology, University Hospitals of Leicester NHS Trust, **59** Viapath, **60** Hub for Biotechnology in the Built Environment, Northumbria University, **61** NU-OMICS Northumbria University, **62** Northumbria University, **63** South Tees Hospitals NHS Foundation Trust, **64** North Cumbria Integrated Care NHS Foundation Trust, **65** North Tees and Hartlepool NHS Foundation Trust, **66** Newcastle Hospitals NHS Foundation Trust, **67** County Durham and Darlington NHS Foundation Trust, **68** Centre for Enzyme Innovation, University of Portsmouth, **69** School of Biological Sciences, University of Portsmouth, **70** Portsmouth Hospitals NHS Trust, **71** University of Warwick, **72** University Hospitals Coventry and Warwickshire, **73** Warwick Medical School and Institute of Precision Diagnostics, Pathology, UHCW NHS Trust, **74** Genomics Innovation Unit, Guy’s and St. Thomas’ NHS Foundation Trust, **75** Centre for Clinical Infection & Diagnostics Research, St. Thomas’ Hospital and Kings College London, **76** Department of Infectious Diseases, King’s College London, **77** Guy’s and St. Thomas’ Hospitals NHS Foundation Trust, **78** Centre for Clinical Infection and Diagnostics Research, Department of Infectious Diseases, Guy’s and St Thomas’ NHS Foundation Trust, **79** Princess Alexandra Hospital Microbiology Dept., **80** Cambridge University Hospitals NHS Foundation Trust, **81** East Kent Hospitals University NHS Foundation Trust, **82** University of Kent, **83** Gloucestershire Hospitals NHS Foundation Trust, **84** Department of Microbiology, Kettering General Hospital, **85** National Infection Service, PHE and Leeds Teaching Hospitals Trust, **86** Cambridge Stem Cell Institute, University of Cambridge, **87** Public Health Scotland, 88 Belfast Health & Social Care Trust, **89** Health Services Laboratories, **90** Barking, Havering and Redbridge University Hospitals NHS Trust, **91** Royal Free NHS Trust, **92** Maidstone and Tunbridge Wells NHS Trust, **93** University of Brighton, **94** Kings College London, **95** PHE Heartlands, **96** Imperial College London, **97** Department of Infection Biology, London School of Hygiene and Tropical Medicine.

## References

1. Kursumovic E, Lennane S, Cook TM. Deaths in healthcare workers due to COVID-19: the need for robust data and analysis. Anaesthesia 2020; 75(8):989–992.

2. Wang D, Hu B, Hu C, Zhu F, Liu X, Zhang J, et al. Clinical Characteristics of 138 Hospitalized Patients With 2019 Novel Coronavirus–Infected Pneumonia in Wuhan, China. JAMA 2020; 323(11):1061–1069.

3. Leclerc Q, Fuller N, Knight L, null n, Funk S, Knight G. What settings have been linked to SARS-CoV-2 transmission clusters? [version 2; peer review: 2 approved]. Wellcome Open Research 2020; 5(83).

4. The DELVE Initiative. Scoping Report on Hospital and Health Care Acquisition of COVID-19 and its Control. DELVE Report No. 3. Published 06 July 2020. In. http://rs-delve.github.io/reports/2020/07/06/nosocomial-scoping-report.html; 2020.

5. Shah ASV, Wood R, Gribben C, Caldwell D, Bishop J, Weir A, et al. Risk of hospital admission with coronavirus disease 2019 in healthcare workers and their households: nationwide linkage cohort study. BMJ 2020; 371:m3582.

6. Lauer SA, Grantz KH, Bi Q, Jones FK, Zheng Q, Meredith HR, et al. The Incubation Period of Coronavirus Disease 2019 (COVID-19) From Publicly Reported Confirmed Cases: Estimation and Application. Annals of Internal Medicine 2020; 172(9):577–582.

7. He X, Lau EHY, Wu P, Deng X, Wang J, Hao X, et al. Temporal dynamics in viral shedding and transmissibility of COVID-19. Nature Medicine 2020; 26(5):672–675.

8. Rivett L, Sridhar S, Sparkes D, Routledge M, Jones NK, Forrest S, et al. Screening of healthcare workers for SARS-CoV-2 highlights the role of asymptomatic carriage in COVID-19 transmission. eLife 2020; 9:e58728.

9. Oran DP, Topol EJ. Prevalence of Asymptomatic SARS-CoV-2 Infection. Annals of Internal Medicine 2020; 173(5):362–367.

10. Lucey M, Macori G, Mullane N, Sutton-Fitzpatrick U, Gonzalez G, Coughlan S, et al. Whole-genome Sequencing to Track Severe Acute Respiratory Syndrome Coronavirus 2 (SARS-CoV-2) Transmission in Nosocomial Outbreaks. Clinical Infectious Diseases 2020.

11. Brown JR, Roy S, Shah D, Williams CA, Williams R, Dunn H, et al. Norovirus Transmission Dynamics in a Pediatric Hospital Using Full Genome Sequences. Clinical Infectious Diseases 2018; 68(2):222–228.

12. Houldcroft CJ, Roy S, Morfopoulou S, Margetts BK, Depledge DP, Cudini J, et al. Use of Whole-Genome Sequencing of Adenovirus in Immunocompromised Pediatric Patients to Identify Nosocomial Transmission and Mixed-Genotype Infection. The Journal of Infectious Diseases 2018; 218(8):1261–1271.

13. Roy S, Hartley J, Dunn H, Williams R, Williams CA, Breuer J. Whole-genome Sequencing Provides Data for Stratifying Infection Prevention and Control Management of Nosocomial Influenza A. Clinical Infectious Diseases 2019; 69(10):1649–1656.

14. Meredith LW, Hamilton WL, Warne B, Houldcroft CJ, Hosmillo M, Jahun AS, et al. Rapid implementation of SARS-CoV-2 sequencing to investigate cases of health-care associated COVID-19: a prospective genomic surveillance study. The Lancet Infectious Diseases 2020; 20(11):1263–1272.

15. Fauver JR, Petrone ME, Hodcroft EB, Shioda K, Ehrlich HY, Watts AG, et al. Coast-to-Coast Spread of SARS-CoV-2 during the Early Epidemic in the United States. Cell 2020; 181(5):990-996.e995.

16. van Dorp L, Acman M, Richard D, Shaw LP, Ford CE, Ormond L, et al. Emergence of genomic diversity and recurrent mutations in SARS-CoV-2. Infection, Genetics and Evolution 2020; 83:104351.

17. The COVID-19 Genomics UK (COG-UK) consortium. An integrated national scale SARS-CoV-2 genomic surveillance network. The Lancet Microbe 2020; 1(3):e99–e100.

18. Public Health England. COVID-19: epidemiological definitions of outbreaks and clusters in particular settings. In. https://www.gov.uk/government/publications/covid-19-epidemiological-definitions-of-outbreaks-and-clusters; 2020.

19. ARTIC Network. SARS-CoV-2 sequencing protocol. In; 2020.

20. Rockett RJ, Arnott A, Lam C, Sadsad R, Timms V, Gray K-A, et al. Revealing COVID-19 transmission in Australia by SARS-CoV-2 genome sequencing and agent-based modeling. Nature Medicine 2020; 26(9):1398–1404.

21. HOCI Sequence Reporting Tool working group. HOCI-COV-GLUE software repository. In; 2020.

22. Singer J, Gifford R, Cotten M, Robertson D. CoV-GLUE: A Web Application for Tracking SARS-CoV-2 Genomic Variation. In: Preprints.org; 2020.

23. Rickman HM, Rampling T, Shaw K, Martinez-Garcia G, Hail L, Coen P, et al. Nosocomial Transmission of Coronavirus Disease 2019: A Retrospective Study of 66 Hospital-acquired Cases in a London Teaching Hospital. Clinical Infectious Diseases 2020.

24. Carter B, Collins JT, Barlow-Pay F, Rickard F, Bruce E, Verduri A, et al. Nosocomial COVID-19 infection: examining the risk of mortality. The COPE-Nosocomial Study (COVID in Older PEople). Journal of Hospital Infection 2020; 106(2):376–384.

25. Houlihan CF, Vora N, Byrne T, Lewer D, Kelly G, Heaney J, et al. Pandemic peak SARS-CoV-2 infection and seroconversion rates in London frontline health-care workers. The Lancet 2020; 396(10246):e6–e7.

26. Volz EM, Frost SDW. Inferring the Source of Transmission with Phylogenetic Data. PLOS Computational Biology 2013; 9(12):e1003397.

27. Villabona-Arenas CJ, Hanage WP, Tully DC. Phylogenetic interpretation during outbreaks requires caution. Nature Microbiology 2020; 5(7):876–877.

